# *MDGA2* homozygous loss-of-function variants cause developmental and epileptic encephalopathy

**DOI:** 10.1101/2025.08.28.25330873

**Authors:** Heba Morsy, Hyeonho Kim, Gyubin Jang, Maha S Zaki, Eleanor Self, Ibrahim Abdelrazek, Mariasavina Severino, Raidah Saleem Albaradie, Khadijah Bakur, Stephanie Efthymiou, Haytham Hussein, Mahmoud M Noureldeen, Bader Alhaddad, Hasnaa M Elbendary, Javeria Raza Alvi, Annarita Scardamaglia, David Murphy, Vicente A Yépez, Julien Gagneur, Tarek I Omar, Marwa Abd Elmaksoud, Jana Vandrovocova, Ebtessam Abdallah, Mary M Reilly, Tipu Sultan, Fowzan S Alkuraya, Joseph G Gleeson, Ji Won Um, Henry Houlden, Jaewon Ko, Reza Maarofian

**Affiliations:** Department of Neuromuscular Diseases, UCL Institute of Neurology, University College London, Queen Square, London WC1N 3BG, UK; Department of Human Genetics, Medical Research Institute, Alexandria University, 21456, Alexandria, Egypt; Department of Brain Sciences, Daegu Gyeongbuk Institute of Science and Technology (DGIST), Daegu 42988, Korea; Center for Synapse Diversity and Specificity, DGIST, Daegu 42988, Korea; Clinical Genetics Department, Human Genetics and Genome Research Institute, National Research Centre, Cairo, Egypt; Neuroradiology Unit, IRCCS Istituto Giannina Gaslini, Genoa, 16147, Italy; King Fahd Specialist Hospital, Dammam, Saudi Arabia; Lifera Omics, Riyadh, 13519, Saudi Arabia; Kuwait Hospital, Sabah Al-Salem, Block1, Kuwait; Department of Paediatrics, Faculty of Medicine, Beni-Suef University, Beni-Suef, Egypt; Department of Paediatric Neurology, Institute of Child Health, Children’s Hospital Lahore, Lahore, Pakistan; Department of Clinical and Movement Neurosciences, UCL Queen Square Institute of Neurology, Queen Square, London WC1N 3BG, UK; School of Computation, Information and Technology, Technical University of Munich, Garching, Germany; Computational Health Centre, Helmholtz Centre Munich, Neuherberg, Germany; Institute of Human Genetics, School of Medicine, Technical University of Munich, Munich, Germany; Neurology Unit, Department of Paediatrics, Faculty of Medicine, Alexandria University, Egypt; Department of Translational Genomics, Genomic Medicine Centre of Excellence, King Faisal Specialist Hospital and Research Centre, Riyadh, 11211, Kingdom of Saudi Arabia; College of Medicine, Alfaisal University, Riyadh, Saudi Arabia; Department of Neurosciences, University of California, San Diego, La Jolla 92093, CA, USA

**Keywords:** MDGA2, neurodevelopmental delay, epileptic encephalopathy, excitatory synapse

## Abstract

*MDGA2* encodes a membrane-associated protein that is critical for regulating glutamatergic synapse development, modulating neuroligins (Nlgns), and maintaining the balance between excitatory and inhibitory synapses. Although MDGA2 has been extensively studied in murine and cellular models, its association with human developmental disorders has not been established to date. Through exome sequencing we identified six distinct homozygous loss-of-function *MDGA2* variants in eight individuals from six consanguineous families, all presenting with developmental and epileptic encephalopathy (DEE). Clinically, these patients exhibited infantile hypotonia, severe neurodevelopmental delay, intractable seizures, progressive brain atrophy, and consistent dysmorphic features, including high anterior hairline, high-arched eyebrows, broad nasal ridge, tented upper lip, and large, low-set ears. Functional studies of three representative nonsense variants in mammalian expression systems and hippocampal cultured neurons revealed impaired MDGA2 membrane trafficking, disrupted Nlgn1 interaction and perturbation of MDGA2-mediated synaptic functions. Altogether, our findings definitively establish *MDGA2* as a novel gene for autosomal recessive DEE subtype, with the pathogenesis explained by loss-of-function mechanism. This discovery underscores the previously unrecognized role of *MDGA2* in human synaptic development and regulation, significantly expanding our understanding of the genetic architecture of DEE.

## Main text

Developmental and epileptic encephalopathies (DEEs) are a group of severe neurodevelopmental disorders characterised by early-onset, intractable seizures and intellectual disability or developmental regression. These conditions have complex and heterogeneous aetiology and usually result from genetic variants that interfere with normal brain development and function. Despite significant advances in the identification of the genetic causes involved in DEEs, many affected individuals remain without defined molecular defects and, hence, adequate genetic counselling. Establishing a genetic basis for these conditions considerably influences treatment strategies and clinical decision-making.^1,2^

In the present study, we identified 16 affected individuals, 8 of them had passed away. Eight affected individuals are included in our cohort, 5 males and 3 females, aged 6 months-17 years, with common clinical features, summarized in **Table 1**.

**Table 1.**
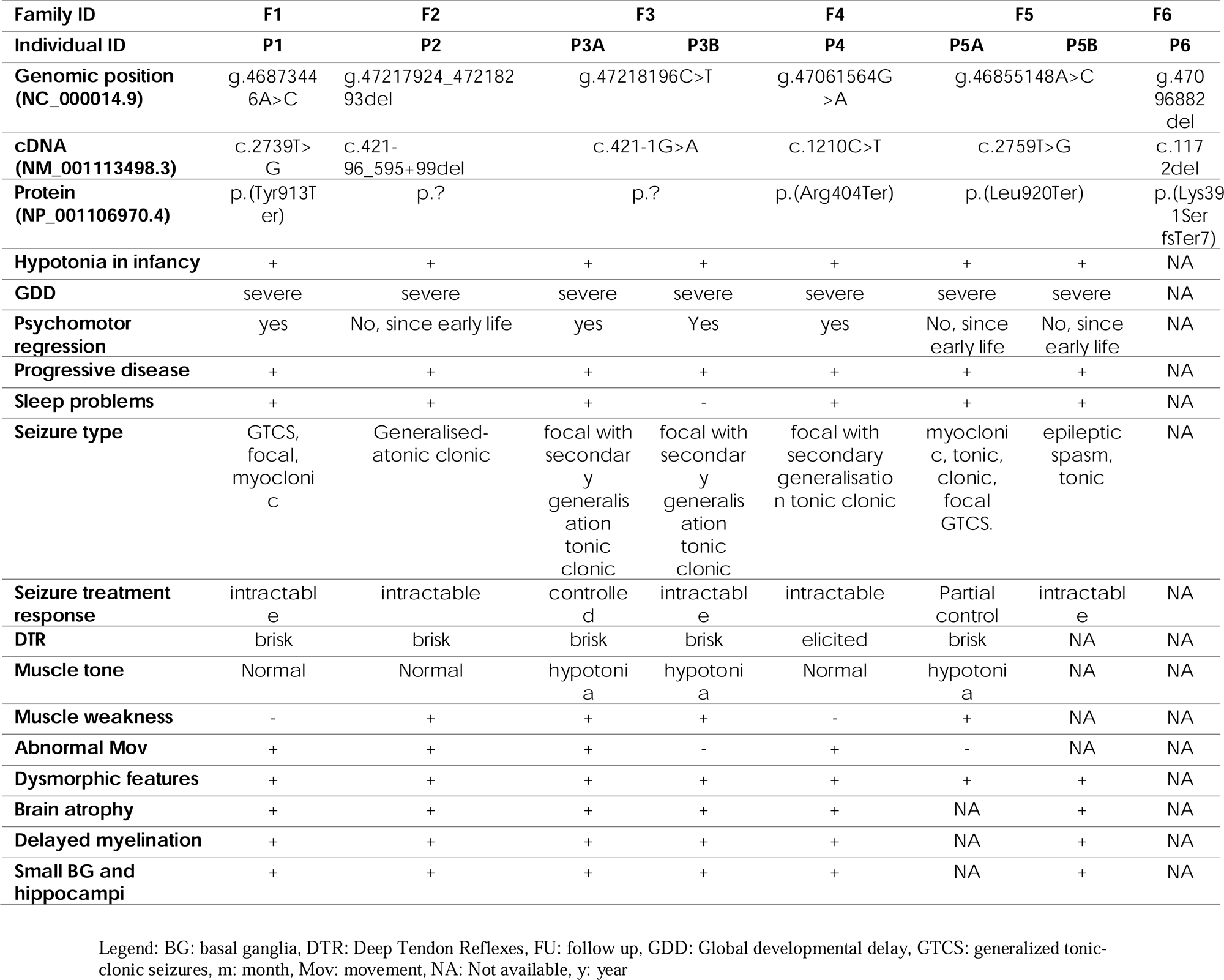
Clinical summary of individuals with homozygous *MDGA2*-related DEE variants.

All individuals presented with DEE and global developmental delay. The ethnicity of all families was Middle Eastern (Egyptians and Saudis) except family 4 which originated from Southeast Asia (Punjabi). All families reported consanguinity, and family histories consistently revealed similarly affected siblings and relatives, often with early mortality. Although a detailed clinical examination was not available for P6, as the patient passed away early in life, we included this individual in the cohort due to the presence of a frameshift variant consistent with the proposed loss-of-function mechanism, along with core clinical features of the disorder, including severe DEE and global developmental delay. We will focus on the description below on the seven patients from families 1-5.

All patients exhibited severe motor and language delay, most failed to reach their main developmental milestones, none achieved independent walking, and only one (P3A) demonstrated transient unsupported sitting. Severe infantile hypotonia was universally observed. Global developmental delay was severe in all cases. Psychomotor regression was reported in four patients, often beginning within the first few months of life. The disease course was progressive in all individuals. Behavioural symptoms were present in four individuals, with sleep disturbances reported in all but one patient (P3B).

Seizures began early in life for all patients and had multiple types including generalized tonic-clonic, focal, myoclonic, tonic, epileptic spasms and atonic seizures. Status epilepticus was recurrent in three patients (P3A, P3B, and P4). Electroencephalogram (EEG) findings showed generalized slowing and encephalopathy in all seven Individuals. Treatment response to anti-epileptic drugs (AEDs) was generally poor, with P4 and P5B achieving some seizure control using a ketogenic diet. Deep-tendon reflexes were brisk in all patients but one (P4), and muscle tone abnormality was noted in P3A, P3B, and P5A who exhibited hypotonia and muscle weakness. No evidence of muscle atrophy or sensory/autonomic abnormality was found. Abnormal movements were observed in all patients except P3B and P5A.

All affected individuals exhibited craniofacial abnormalities, with some degree of frontal prominence observed in all cases. A high anterior hairline and high forehead were consistent findings across most individuals (6/7). Dolichocephaly was observed in P1 and P3A, while intertemporal narrowing was shared by P1, P3B, and P5A. Periorbital features included thin, highly arched eyebrows, noted in nearly all individuals (6/7), occasionally with a medial flare (P3A). Strabismus along with upslanting palpebral fissures was a frequent finding in our cohort (observed in 5/7 and 4/7, respectively). Long eyelashes were observed in P2, P3B, and P5B. The nasal region was consistently affected, with all individuals presenting a broad nasal bridge. Additional features such as underdeveloped nasal alae and bulbous nasal tip were variably present. Orofacial abnormalities were prominent, with a tented upper lip observed in almost all cases. In the affected siblings in family 5, this was accompanied by a thin lower lip. Downturned corners of the mouth were noted in P4 and P5B. Most of individuals had large, low-set ears. An age-related progression in dysmorphic features was apparent, with facial characteristics becoming more distinct over time.

Brain MRI studies were available for review in 6/8 subjects and were performed at an average age of 1.2 years (range 6 months – 3 years). In all cases there was delayed/incomplete myelination and early onset brain atrophy with thinning of the white matter, basal ganglia volume loss, and small hippocampi (**Figure 1B**).

**Figure 1.**
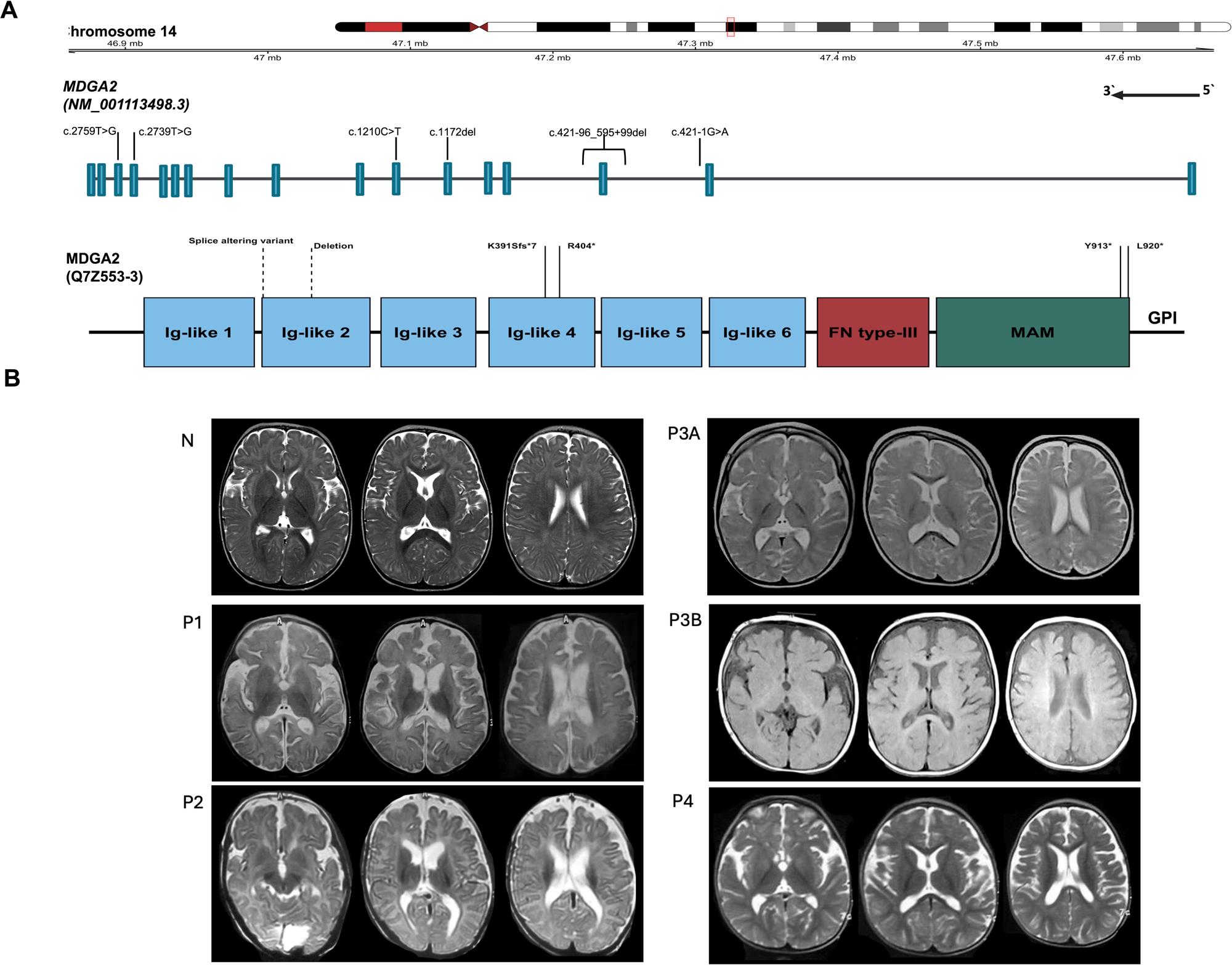
Genetic variants, and neuroradiological findings of individuals with *MDGA2*-related DEE variants. (**A**) Schematic structure of *MDGA2* gene (top) and its coding protein (bottom) highlighting variants identified in this study. Deletion and splice variants are represented in the MDGA2 protein approximately by dashed lines, as exact positions in protein are not known. (**C**) Neuroradiological features. Brain MRI of a normal control (N) before 1 year, of subjects P1, P2, P3b before 1 year, and subjects P3a and P4. All subjects show delayed or incomplete myelination, associated with moderate-to-severe white matter volume loss and consequent ventricular dilatation. Increased cerebral subarachnoid spaces and reduced basal ganglia volume, particularly in the caudate nuclei and thalami, are also observed.

Ethical approvals were obtained in each recruiting centre by the appropriate institutional review board (IRB), and all participants parents provided informed consent to participate in the research study. Proband-only whole exome sequencing (WES) revealed six homozygous variants in *MDGA2* gene (MIM: 611128) in eight affected individuals. All identified variants were absent from gnomAD v3^3^ and lie within a region of homozygosity (ROH). (**Tables S1–S8, Figure S1**). Segregation of SNVs was confirmed by Sanger sequencing in available family members. (**Figure S2).**

The homozygous *MDGA2-*related DEE variants (GenBank: NM_001113498.3 and NP_001106970.4) identified include three nonsense variants (p.Tyr913Ter in P1, p.Arg404Ter in P4, and p.Leu920Ter in P5A and P5B), one splice variant (c.421-1G>A in P3A and P3B), one frameshift variant (p.Lys391SerfsTer7 in P6) and a homozygous deletion of exon 3 (c.421-96_595+99del) in P2 (**Figure 1A, Figures S3-S5**). All identified variants are predicted to result in loss-of-function (LoF), which supports their pathogenicity considering *MDGA2*’s intolerance to loss-of-function variants.^3^ The three nonsense variants are not in the last or penultimate exon of *MDGA2,* suggesting that mutated mRNA product could have increased susceptibility to nonsense-mediated mRNA decay (NMD). The splice variant identified in family 3 has a SpliceAI^4^ delta score of 0.99 and is predicted to cause loss of the acceptor splice site; it is likely to give rise to abnormal spicing and therefore a loss-of-function effect. In addition, testing the splicing effect of this variant using AbSplice^5^ showed a score of 0.054 (greater than the medium cutoff of 0.05) in multiple Brain tissue subtypes. The 370-bp homozygous deletion identified in P2 (NC_000014.9: g.47217924_47218293del) contains all of exon 3, and is predicted to lead to a frameshift, and the resulting mRNA is thus likely to undergo NMD. The NP_001106970.4: p.(Lys391SerfsTer7) variant; introduces a frameshift at codon 391, resulting in a premature stop codon. This is predicted to produce a truncated MDGA2 protein or trigger NMD. The six *MDGA2* variants reported herein might cause pathological truncation of MDGA2 protein, decrease *MDGA2* mRNA stability, and/or increase its degradation by NMD, as reported in other human genetic diseases.^6^

*MDGA2* is an evolutionarily conserved gene on chromosome 14q21.3. MDGA2, and its paralog MDGA1, are one of the neuronal glycosylphosphatidylinositol (GPI) - anchored proteins. Both are membrane associated proteins that contain six tandem immunoglobulin (Ig)-like domains, a fibronectin-like region, a single meprin, A-5 protein, and receptor protein-tyrosine phosphatase mu (MAM) domain, and a C-terminal GPI anchor. They have been highlighted as key suppressive factors that tune the balanced activity of neural circuits. MDGAs proteins expression is restricted to the central nervous system, begins early in development, and continues throughout adulthood.^7–12^

Prior studies established that MDGA2 negatively modulates glutamatergic synapses via distinct extracellular mechanisms. Studies using knockout (KO) mice showed that Mdgas genes dysregulation could be associated with a subset of neuropsychiatric disorders, such as autism spectrum disorders (ASDs) and schizophrenia. It has been shown that mutation of Mdga2 elevates excitatory transmission, and Mdga2 modulates neuroligin-1 interaction with neurexins and suppresses excitatory synapse development.^9,13–17^ To explore the effects of *MDGA2* variants on protein function, we tested three representative *MDGA2* nonsense variants; Y844*, R335* and L851*, all described based on the canonical protein isoform Q7Z553-1. (**Figures S6A and S7A**) The Y844* nonsense variant corresponds to the variant identified in individual P1 (Y913*) in the Q7Z553-3 isoform. Notably, these residues are evolutionarily conserved across species, hinting at their possible functional significance (**Figure S7B**). We assessed the impact of three nonsense *MDGA2* variants using a mammalian cDNA expression vector but could not express cDNAs for the splice or deletion variants.

We first examined the expression levels and intracellular transport properties of the *MDGA2* variants upon expression in human embryonic kidney 293T (HEK293T) cells. HEK293T cells were transfected with vectors encoding hemagglutinin (HA)-tagged full-length MDGA2 wild-type (WT) or variants. Immunoblotting of cell lysates showed that the total protein expression levels of the MDGA2 Y844* and L851* were comparable to those of MDGA2 WT, whereas the R335* variant was not (**Figure S6B**). All three MDGA2 nonsense variants were expected to yield truncated proteins because they lacked membrane-anchored GPI sequence. However, whereas expressed Y844* and L851* migrated to slightly lower positions on SDS-PAGE analyses and were not secreted, the R335* variant, yielded a truncated protein that was ∼40 kDa on SDS-PAGE analyses and was prominently secreted (**Figure S6B**). We next examined the surface and intracellular protein levels of WT and variant MDGA2 proteins in HEK293T cells (**Figures S6C** and **S6D**). None of the tested MDGA2 nonsense variants displayed detectable surface expressions in HEK293T cells, possibly reflecting their complete entrapment in an intracellular compartment.

To determine whether these three MDGA2 nonsense variants would alter the interactions with neuroligin-1 (Nlgn1), an extracellular ligand of MDGA2, we assayed the cell-surface binding of recombinant Ig-fusion proteins of Nlgn1 (Ig-Nlgn1) or IgC alone (negative control) with HEK293T cells expressing HA-tagged MDGA2 variants (**Figures S6E** and **S6F**). Our results revealed that the IgC-Nlgn1 proteins robustly bound to HEK293T cells expressing MDGA2 WT, but not the surface transport-defective MDGA2 variants. IgC did not bind to any tested MDGA2 variant. Overall, our results suggest that the tested MDGA2 nonsense variants lack the surface-trafficking and ligand-binding activities exhibited by MDGA2 WT.

MDGA2 was previously shown to inhibit the Nlgn1-mediated synaptogenic activity in driving presynaptic assembly in heterologous synapse formation assays, where Nlgn1 expressed in heterologous cells recruits presynaptic components in axons of cocultured neurons.^12^ To test whether these three nonsense MDGA2 variants (R335*, Y844*, and L851*) affected the ability of MDGA2 WT to inhibit Nlgn1-induced presynaptic differentiation-inducing activity, we performed heterologous synapse-formation assays using HEK293T cells expressing EGFP alone (negative control), expressing Nlgn1 alone, or coexpressing Nlgn1 with the indicated MDGA2 variants, and applying anti-synapsin antibodies to label presynaptic sites **(Figure 2A**). Nlgn1 expressed in HEK293T cells strongly recruited presynaptic synapsin puncta in axons of cocultured neurons, and coexpressed MDGA2 WT effectively decreased Nlgn1-induced presynaptic differentiation (**Figures 2B** and **2C**). In contrast, none of the MDGA2 nonsense variants inhibited Nlgn1-mediated synaptogenic activity (**Figures 2B** and **2C**). These observations are consistent with the above-described findings that all three nonsense MDGA2 variants lacked proper surface transport and ligand-binding activities.

**Figure 2.**
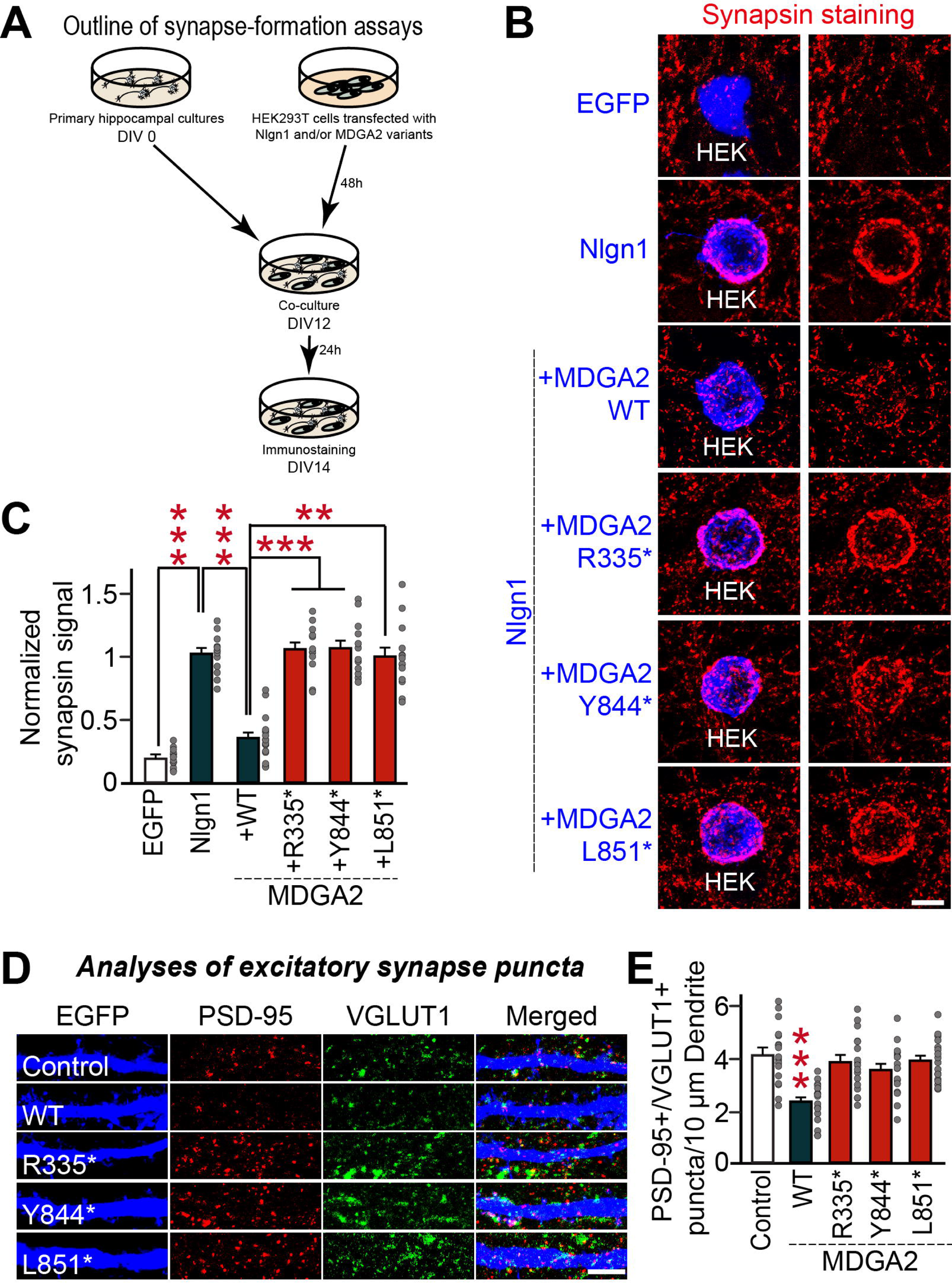
Nonsense *MDGA2* variants exhibit inactivation of anti-synaptogenic activities and impairment of excitatory synapse suppression ability in regulating synapse numbers in cultured hippocampal neurons. (**A**) Overview of heterologous synapse formation assays performed in the current study. (**B** and **C**) Representative images (**B**) and summary graphs (**C**) showing the density of synaptic puncta of cultured hippocampal neurons transfected at DIV7 with the indicated full-length MDGA2 expression construct and immunostained at DIV14 with antibodies to VGLUT1/PSD-95 and EGFP. Data are presented as means ± SEMs (\*\*\**p*L<L0.001; ANOVA with non-parametric Kruskal–Wallis test; n = 16–17 images/group). (**D** and **E**) Effects of MDGA2 WT or variants on Nlgn1 synaptogenic activities. HEK293T cells expressing the indicated proteins were cocultured with hippocampal neurons without or with coexpression of the indicated version of MDGA2. Representative images (**D**) of cocultures immunostained with antibodies to EGFP or HA (blue) and synapsin I (red). Quantitation (**E**) of heterologous synapse-formation assay results, as determined by calculating the signal ratio of synapsin to EGFP/HA. Data are presented as means ± SEMs (\*\**p* < 0.01, \*\*\**p*L<L0.001; nonparametric Kruskal–Wallis test with Dunn’s *post-hoc* test; n = 12–13 cells/group). Scale bar, 10 μm (applies to all images).

We next examined whether the MDGA2 variants influenced the ability of MDGA2 WT to suppress the number of excitatory synapses in cultured hippocampal neurons. To this end, we co-transfected cells with expression vectors encoding EGFP and the various versions of MDGA2 at DIV7, and immunostained the transfected neurons with antibodies to VGLUT1 (a marker for excitatory presynaptic terminal), PSD-95 (a marker for excitatory postsynaptic density), and EGFP (to visualize the transfected neurons) at DIV14. The density of excitatory synaptic puncta immunoreactive to both VGLUT1 and PSD-95 was significantly decreased in MDGA2 WT-expressing neurons (**Figures 2D** and **2E**), in line with our recent report. ^13^ In contrast, overexpression of all three MDGA2 nonsense variants failed to alter the density of excitatory synaptic puncta, in agreement with our results showing that these variants have deficits in surface transport and synaptogenic activity. Our immunocytochemical analyses indicate that all tested MDGA2 nonsense variants act as loss-of-function variants.

To corroborate these results, we measured miniature excitatory postsynaptic currents (mEPSCs) in cultured hippocampal neurons using whole-cell electrophysiological recordings (**Figure 3A**). We found that the frequency, but not amplitude, of mEPSCs, was markedly decreased in MDGA2 WT-expressing cultured hippocampal neurons (**Figures 3B–3F**), which was in line with our previous results.^13^ Overexpression of each nonsense *MDGA2* variants, failed to alter excitatory synaptic transmission (**Figures 3B–3F**). We next monitored evoked EPSCs by measuring AMPA receptor- and NMDA receptor-mediated EPSCs (AMPA-eEPSCs and NMDA-eEPSCs, respectively) at holding potentials of -70 mV and +40 mV, respectively, in the presence of external Mg^2+^ (**Figure 3A**). Overexpression of MDGA2 WT decreased the amplitude of both AMPA-eEPSCs and NMDA-eEPSCs, as previously reported.^13^ However, overexpression of the *MDGA2* nonsense variant did not alter the amplitude of AMPA-eEPSCs or NMDA-eEPSCs (**Figures 3G–3L**). These results collectively reinforce that all three *MDGA2* nonsense variants exhibit perturbation of MDGA2-mediated synaptic functions.

**Figure 3.**
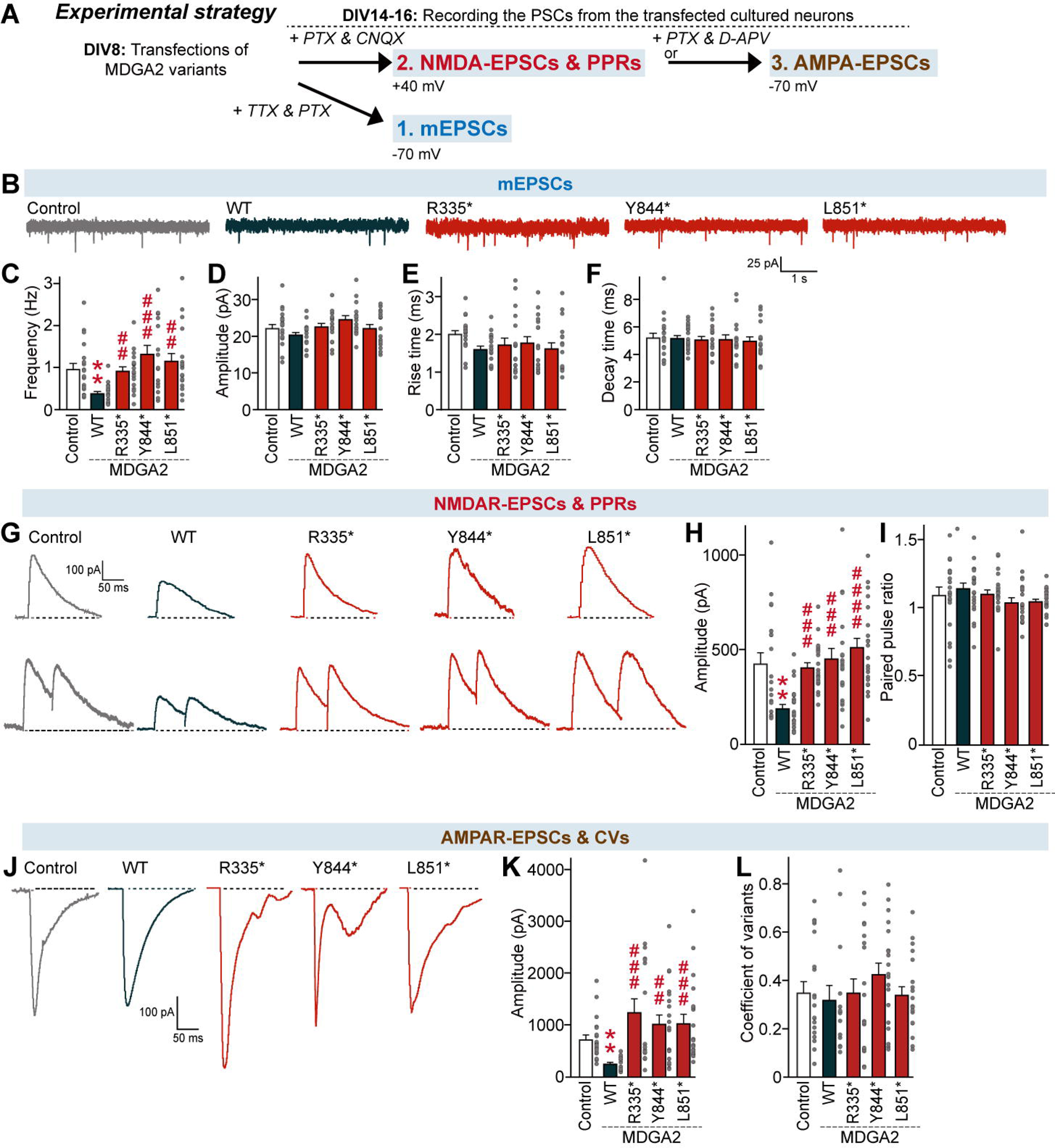
Nonsense *MDGA2* variants exhibit impaired excitatory synapse-suppression activities in regulating basal synaptic transmission of cultured hippocampal neurons. (**A**) Experimental strategy for recording miniature excitatory postsynaptic currents (mEPSCs), evoked AMPAR-mediated EPSCs, and evoked NMDAR-mediated EPSCs from cultured hippocampal neurons transfected with the indicated MDGA2 variants. (**B**–**F**) Representative mEPSC traces (**B**) and quantification of the frequency (**C**), amplitude (**D**), rise time (**E**), and decay time (**F**) of mEPSCs. Data are presented as means ± SEMs (Control, n = 21; MDGA2 WT n = 18; MDGA2 R335*, n = 19; MDGA2 Y844*, n = 17; MDGA2 L851*, n = 18; Control vs. MDGA2: \*\**p* < 0.01, ^##^*p* < 0.01, ^###^*p* < 0.001; ANOVA with a non-parametric Kruskal–Wallis test). (**G**–**L**) Representative evoked NMDAR-EPSCs and NMDAR paired-pulse traces (**G**) and quantification of the amplitude (**H**) and paired pulse ratio (**I**) of evoked NMDAR-EPSCs. Data are presented as means ± SEMs (Control, n = 19; MDGA2 WT n = 21; MDGA2 R335*, n = 20; MDGA2 Y844*, n = 20; MDGA2 L851*, n = 21; \*\**p* < 0.01, ^##^*p* < 0.01, ^###^*p* < 0.001; ANOVA with a non-parametric Kruskal–Wallis test). (**J**–**L**) Representative evoked AMPAR-EPSC traces (**J**) and quantification of amplitude (**K**) and coefficient of variation. (**L**). Data are presented as means ± SEMs (Control, n = 18; MDGA2 WT, n = 13; MDGA2 R335*, n = 17; MDGA2 Y844*, n = 18; MDGA2 L851*, n = 18; \*\**p* < 0.01, ^##^*p* < 0.01, ^###^*p* < 0.001; ANOVA with a non-parametric Kruskal–Wallis test).

Altogether, we describe a novel autosomal recessive DEE syndrome caused by homozygous loss-of-function (LoF) variants in *MDGA2*, identified in eight individuals from six consanguineous families. All individuals exhibited severe developmental delay, intractable seizures, and age-progressive dysmorphic facial features (high-arched eyebrows, broad nasal ridge, tented upper lip, and large, low-set ears). Notably, the condition exhibits high lethality, with many affected individuals dying in early infancy. In mouse models, full knockout of Mdga2 proved to be perinatally lethal, underscoring its critical role in normal brain development.^17^ Neuroimaging revealed nonspecific but consistent findings of early-onset brain atrophy, with volume reduction of the basal ganglia and hippocampi and delayed myelination, similar to findings in STXBP1- and FRRS1L-related DEEs,^18^ marking MDGA2 as a new player in synaptic encephalopathies.

MDGA2, a synaptic regulator, suppresses density, transmission, and strength of glutamatergic synapses, including both NMDA- and AMPA-receptor-mediated postsynaptic responses. It exerts these effects by modulating NLGN1 through its immunoglobulin-like and MAM domains, thereby inhibiting the NLGN1–NEUREXIN (NRXN1) interaction critical for excitatory synapse formation.^13^ Both *NLGN1* and *NRXN1* variants have been linked to neurodevelopmental disorders.^19,20^ Our functional studies demonstrate that homozygous *MDGA2* nonsense variants affecting these key domains, abolish MDGA2’s surface trafficking and NLGN1-binding activity, leading to unchecked excitatory synapse formation. This synaptic dysregulation likely disrupts the cortical excitation–inhibition balance, a known driver of epileptogenesis shared with other DEEs.^21^ Furthermore, the progressive brain atrophy and delayed myelination observed on MRI suggest that MDGA2 LoF exacerbates neuronal loss, potentially amplifying seizure severity.

Beyond seizures, MDGA2 dysfunction contributes to broader neurodevelopmental impairments in murine models, where *Mdga2^+/-^* mice haploinsufficiency causes delayed motor development, reduced ultrasonic vocalizations, and autism-like behaviors—including stereotypies, impaired social interactions, and memory deficits,^17^ paralleling the severe psychomotor delay and behavioural symptoms in our cohort. Additionally, the principal synaptic target for Mdga2, Nlgn1, maintains synchronous cortical activity during wakefulness and sleep,^20^ suggesting that disrupted MDGA2–NLGN1 interactions may underlie the sleep disturbances consistently reported in our patients. These findings establish *MDGA2* LoF as a novel cause of DEE, with its synaptic defects directly contributing to the intractable seizure phenotype and broader neurodevelopmental impairments. The partial seizure control achieved with the ketogenic diet in patients P4 and P5B is particularly noteworthy. This dietary intervention, known to modulate neurotransmitter function, ^22^ lends further support to the hypothesis of excitatory dysregulation in *MDGA2*-related DEE, potentially restoring the excitation-inhibition balance disrupted by MDGA2 dysfunction. While this observation warrants further investigation through clinical trials, understanding the underlying pathogenic mechanism is a crucial first step toward identifying effective therapeutic strategies.

While we tested three representative nonsense variants, the splice, frameshift, and deletion variants are predicted to cause LOF via NMD or protein truncation, consistent with MDGA2’s high LoF intolerance (pLI = 1).^3^ Future studies investigating the pathogenicity of missense variants are worthwhile to examine whether these changes dysregulate the MDGA2-mediated suppression of glutamatergic synapses.

Although MDGA2’s roles have been explored in murine and cellular models, ^13,16,17^ the current study is the first to demonstrate its essential role in human neurodevelopmental disorders. Nevertheless, our functional validation was limited to in-vitro models, due to very low expression of *MDGA2* in blood and skin based on Genotype-Tissue Expression consortium (GTEx) data,^23^ rendering studying *MDGA2*-related DEE variants in peripheral patient-derived samples challenging. This is further elucidated by RNA-sequencing data from blood and fibroblasts in the Solve-RD cohort, where *MDGA2* expression was too low to be properly modelled (**Figure S8**). In contrast, brain-specific gene expression data shows enrichment of MDGA2 in excitatory and inhibitory neurons and oligodendrocyte precursor cells, suggesting it has cell-type-specific roles in the CNS.^24^ In addition, given that MDGA2 is also expressed in astrocytes, which plays an essential role at the glutamatergic synapse,^25^ it would be interesting to apply systematic functional analyses to examine if any aspects of astrocytic functions are altered. Since developing functional models remains challenging due to the gene’s high conservation across species^26^ and low peripheral expression, future studies using patient-derived induced pluripotent stem cell (iPSC) neurons are highly recommended to confirm MDGA2’s role in human synaptic dysfunction, complementing our in-vitro findings.

In conclusion, we establish *MDGA2* as a novel DEE gene, linking its LoF variants to a severe neurodevelopmental syndrome, through integrated clinical, genetic, and functional analyses. This study expands the genetic aetiology of DEEs, holds promise for improving patients’ outcomes with early diagnosis and genetic counselling, and lays the basis for targeted therapies, advancing our understanding of MDGA2 synaptic dysfunction in DEEs.

## Data availability

The data that support the findings of this study are available within the paper and in the supplemental information. Whole exome sequencing data are not publicly available due to privacy or ethical restrictions.

## Acknowledgements

We are grateful for the important support from patients and their families, our UK, international collaborators and SYNAPS Study Group. We are grateful to Jinha Kim (DGIST) for technical assistance. H.M. was supported by the Wellcome Trust grant 220906/Z/20/Z and UCL Global Engagement Fund scheme 2023. H.K. was supported by the National Research Foundation of Korea (NRF) funded by the Ministry of Science and Future Planning (RS-2024-00339642). J.W.U. was supported by the NRF funded by the Ministry of Science and Future Planning (RS-2023-NR076948). J.K. was supported by the NRF funded by the Ministry of Science and Future Planning (RS-2022-NR070708). M.Z. was funded by GERF-STDF: 33650, STDF, Egypt. H.H. was funded by the Wellcome Trust, MRC, MSA Trust, National Institute for Health Research University College London Hospitals Biomedical Research Centre (NIHR-BRC), Michael J Fox Foundation (MJFF), Fidelity Trust, Rosetrees Trust, Dolby Family Fund, Alzheimer’s Research UK (ARUK), MSA Coalition, Parkinson’s Disease Society, Parkinson’s Foundation, Guarantors of Brain, Cerebral Palsy Alliance, FARA, EAN, Victoria Brain Bank, NIH NeuroBioBank, Queen Square BrainBank, and MRC Brainbank Network. V.A.Y. and J.G. were funded by the Deutsche Forschungsgemeinschaft (DFG, German Research Foundation) via the project NFDI 1/1 “GHGA - German Human Genome-Phenome Archive” (#441914366). ERDERA has received funding from the European Union’s Horizon Europe research and innovation programme under grant agreement N°101156595. The TUM IT infrastructure was co-funded by the Deutsche Forschungsgemeinschaft (DFG, German Research Foundation) – Project-ID 461264291

## Author contributions

**Conceptualization,** H.M., R.M., J.K.; **data curation,** H.M., H.K., G.J., D.M., J.W.U., **formal analysis,** H.M, B.A., V.A.Y, J.G., J.V.; **methodology,** H.M., H.K., G.J., E.S., A.S, **funding acquisition,** M.R.R., H.H., J.K. ; **investigation,** all authors, **recruitment and clinical and diagnostic evaluations,** H.M., M.Z., I.A., R.S.A, K.B.,S.E., H. Hussein, M.M.N., H.M.E., J.R.A., T.I.O., M.A., E.A., T.S., F.S.A., H.H; **Supervision,** M.R.R., F.S.A, J.G.G, J.W.U., H.H., J.K., R.M.; **writing – original draft,** H.M., H.K., G.J., M.S., R.M., J.K.; **writing – review and editing,** all authors.

## Web resources

OMIM, https://www.omim.org/

gnomAD, https://gnomad.broadinstitute.org/

GTEX, https://www.gtexportal.org/home/

