## supplemental data for "*MDGA2* homozygous loss-of-function variants cause developmental and epileptic encephalopathy"

**Supplemental figures and legends**

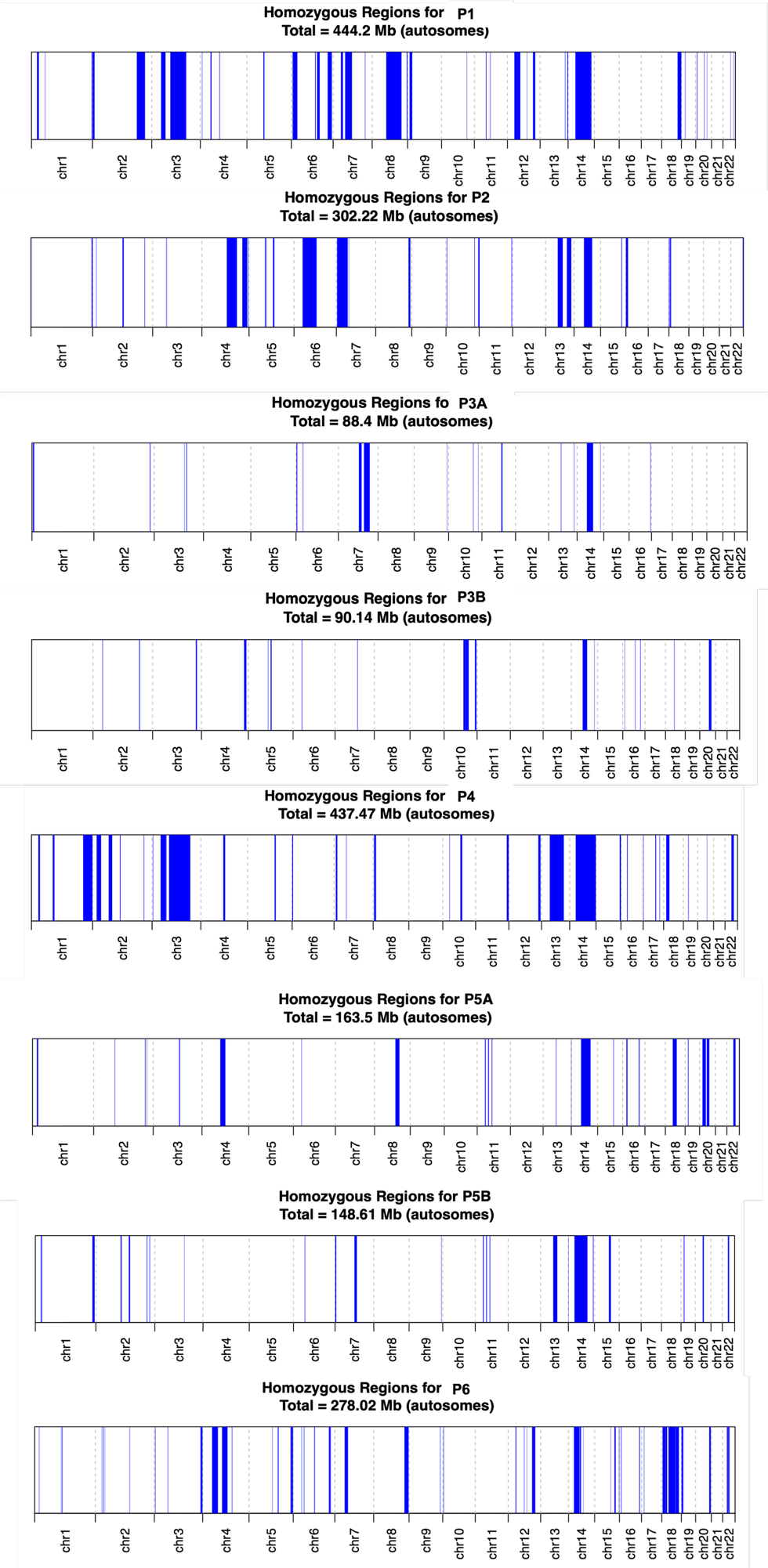

**Figure S2. Genome maps of ROH** generated using Automap v1.0. for the eight individuals included in the study.

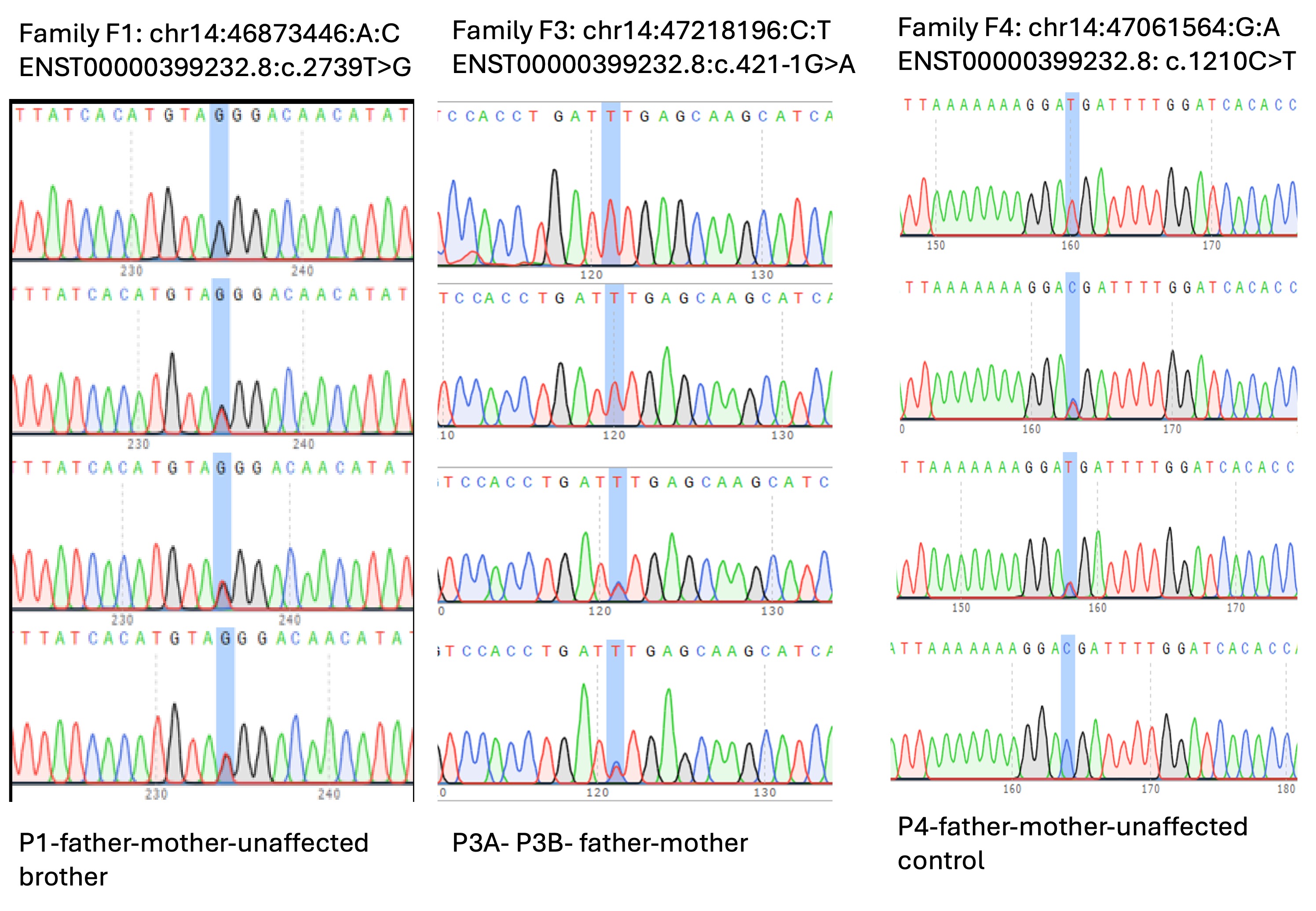

**Figure S3. Chromatograms from Sanger sequencing of *MDGA2* genotypes** for probands, parents, and available siblings of families F1, F3 and F4.

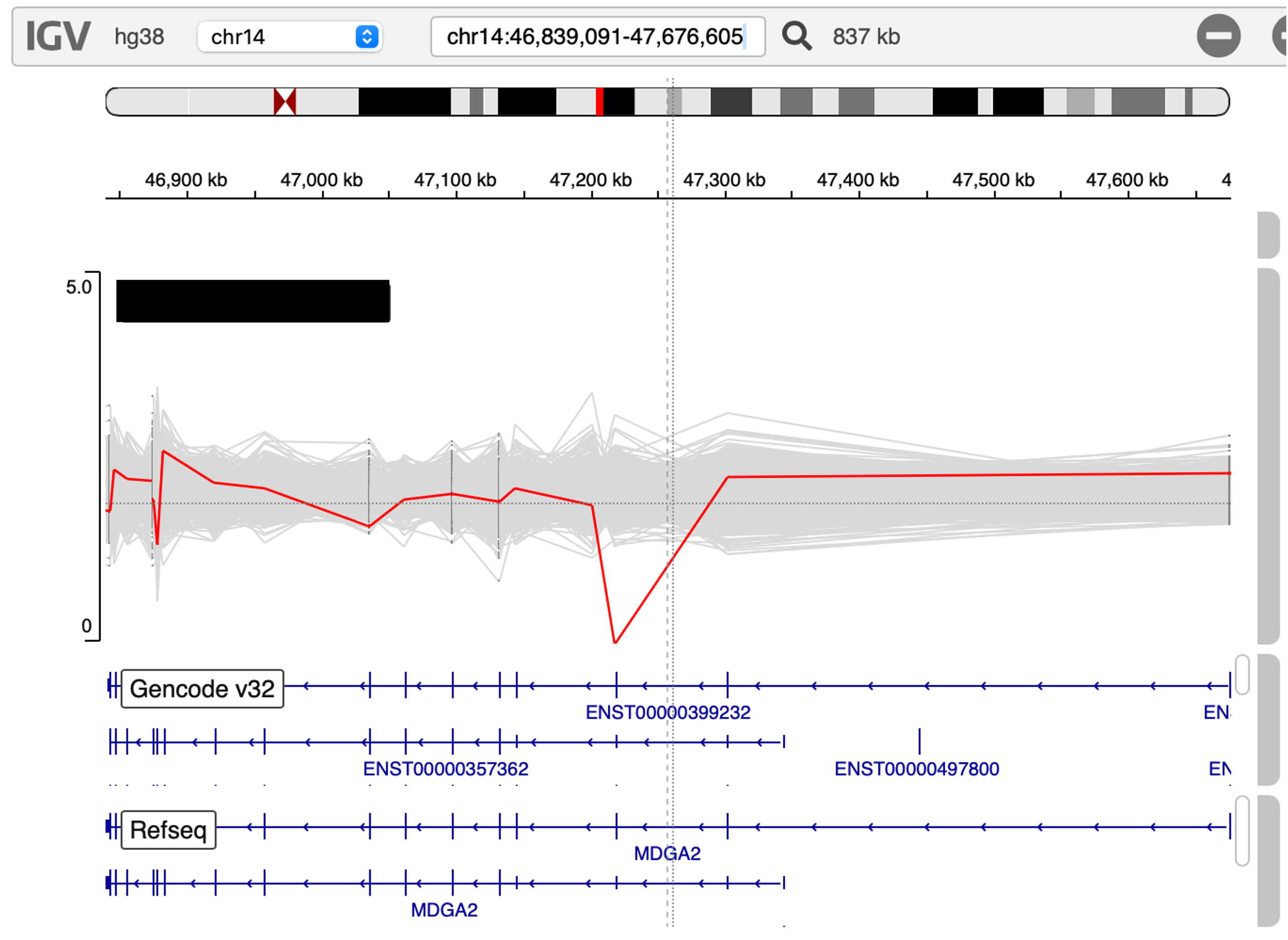

**Figure S4.**  **IGV Screenshot of the coverage for individual P2.** The proband carries a homozygous deletion (Chr14:47217924-47218293: DEL) containing all of exon 3 in *MDGA2.*

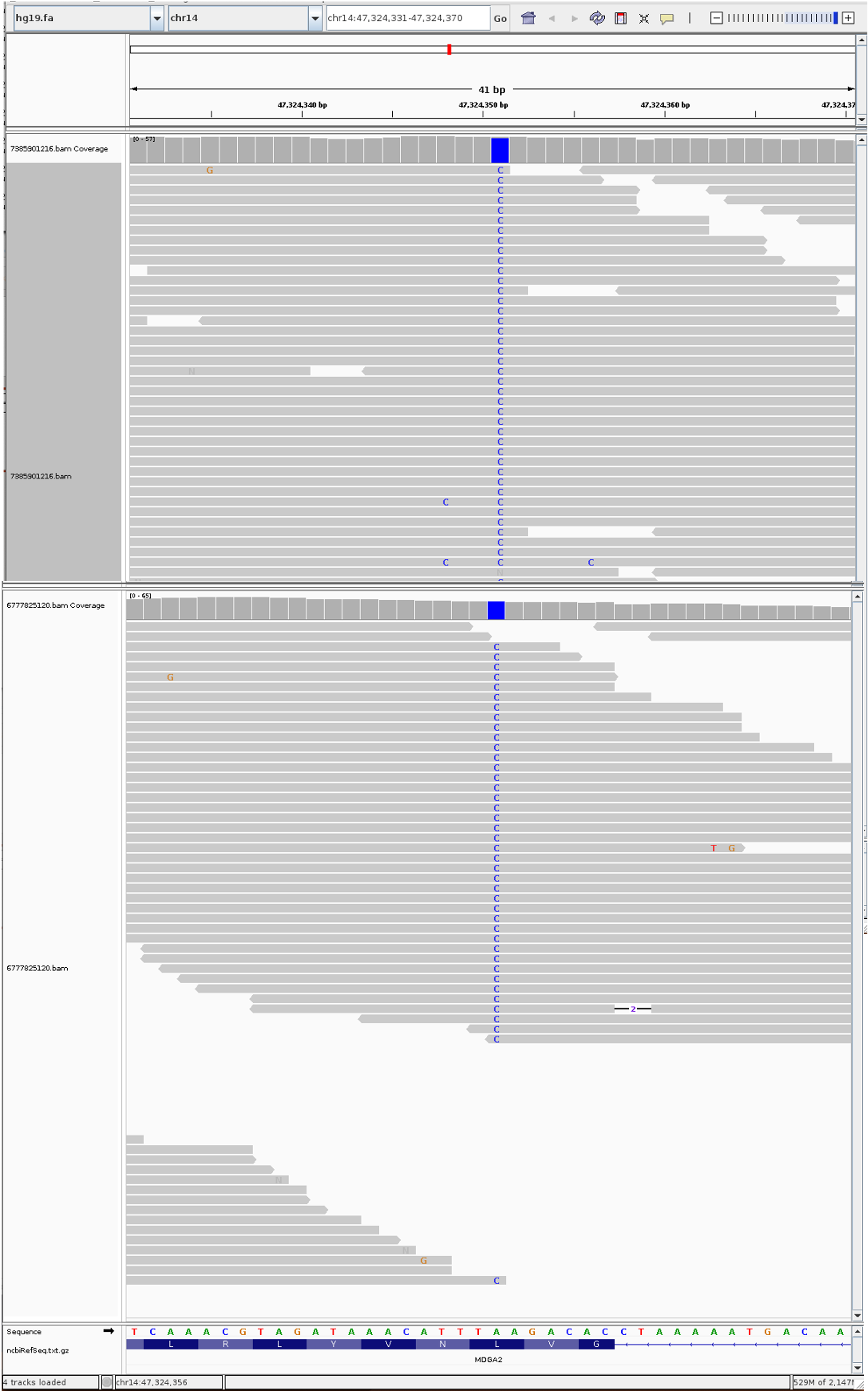

**Figure S5.**  **IGV Screenshot of SNV identified in individuals P5A (top) and P5B (bottom)**

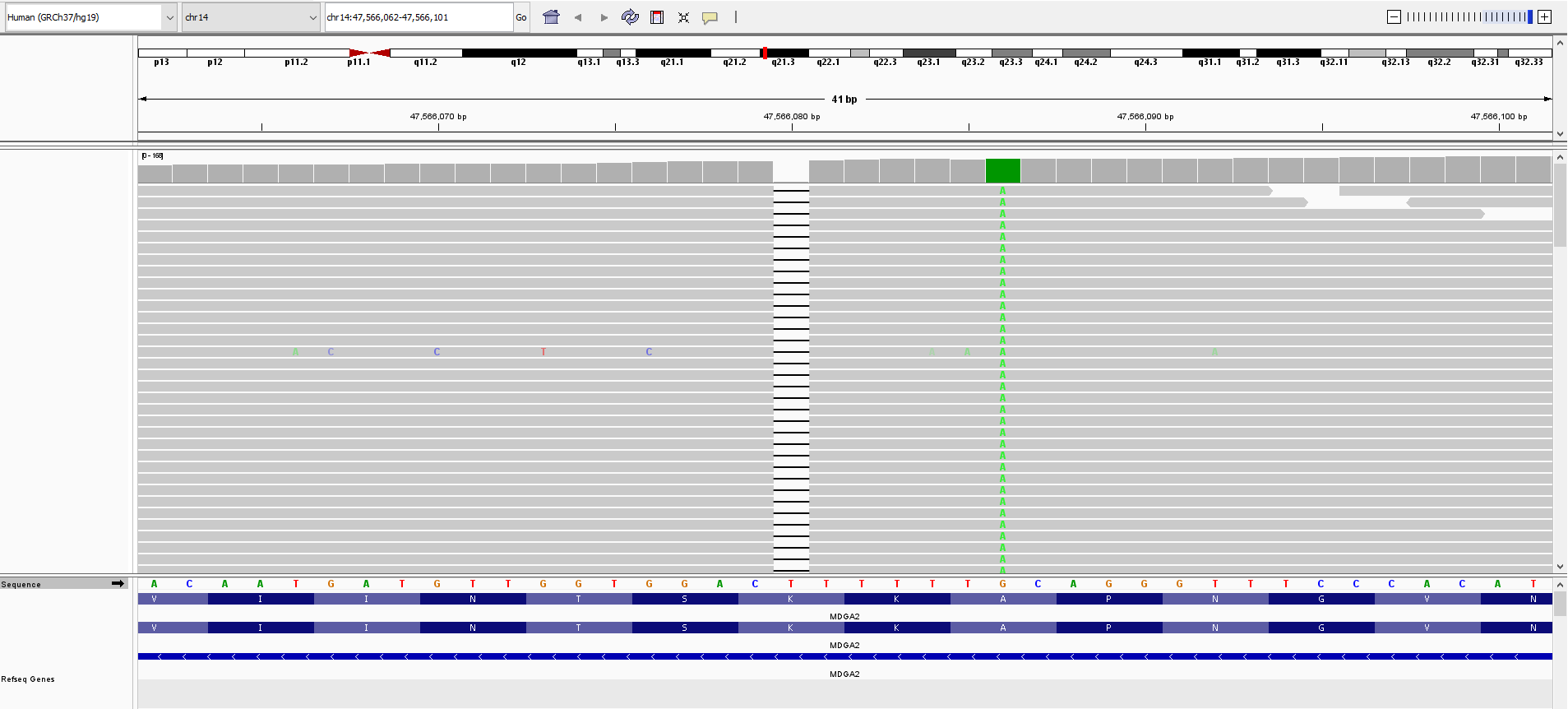

**Figure S6.** **IGV Screenshot of homozygous frameshift deletion identified in individuals P6.**

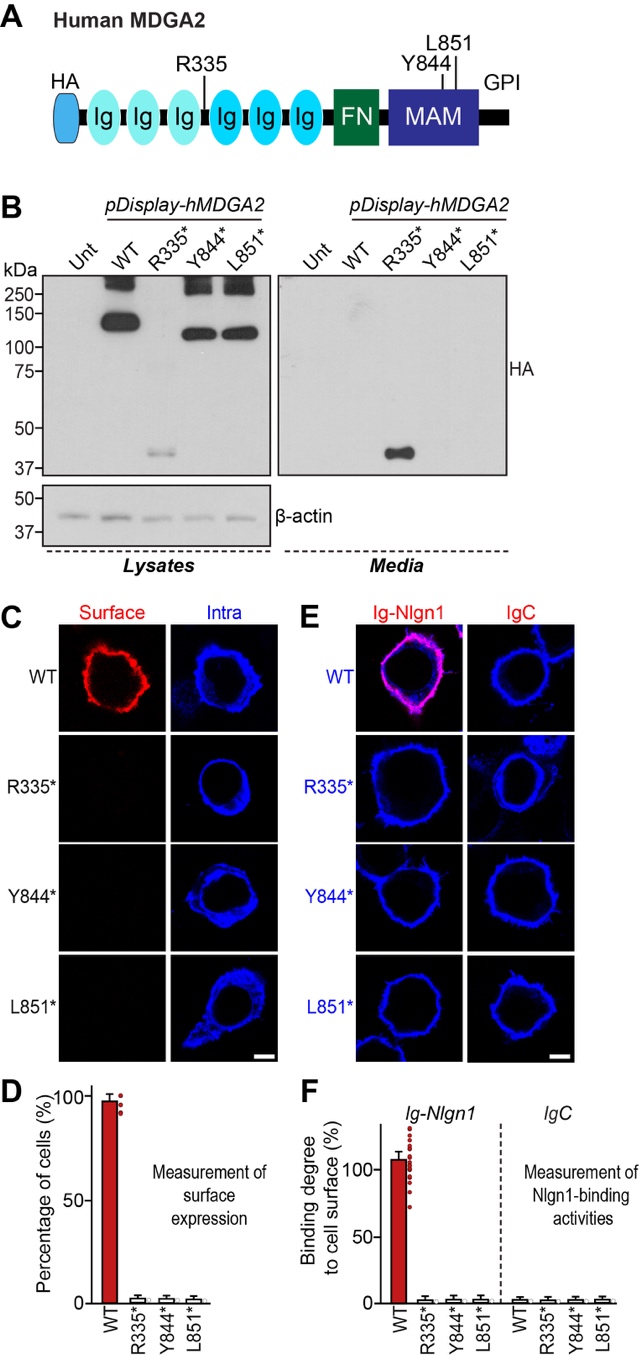

**Figure S7. Tested nonsense MDGA2 variants exhibit impaired surface transport properties in heterologous cells.**

(**A**) Diagram illustrating the MDGA2 variants used in the molecular replacement experiments.

(**B**) Immunoblot analyses probing the expression levels of MDGA2 WT and the indicated variants in HEK293T cells.

(**C** and **D**) Cell surface-binding assays. Representative images (**C**) showing HEK293T cells expressing N-terminally HA-tagged MDGA2 WT or variants incubated with purified Ig-fused neuroligin-1 (Ig-Nlgn1) or negative control (IgC), as analyzed by immunofluorescence imaging for Ig-fusion proteins (red) and HA (green). Quantification of cell surface binding (**D**). Data are shown as means ± SEMs (n = 14–20 cells/group).

(**E** and **F**) Analyses of the surface transport activity of HA-MDGA WT and variants in HEK293T cells. Representative images (**E**) of transfected HEK293T cells illustrating the expression and surface transport activity of MDGA2. Transfected cells were fixed (not permeabilized) and incubated with anti-HA antibody to detect the extracellular region of the MDGA2 (red). The intracellular level of each MDGA2 construct was visualized by incubating permeabilized cells with an anti-HA antibody (green). Quantification (**F**) of the surface transport activity of MDGA2 constructs. Data are presented as means ± SEMs (nonparametric Kruskal–Wallis test with Dunn’s *post-hoc* test; n = 8 images/group).

Scale bar, 10 μm (applies to all images).

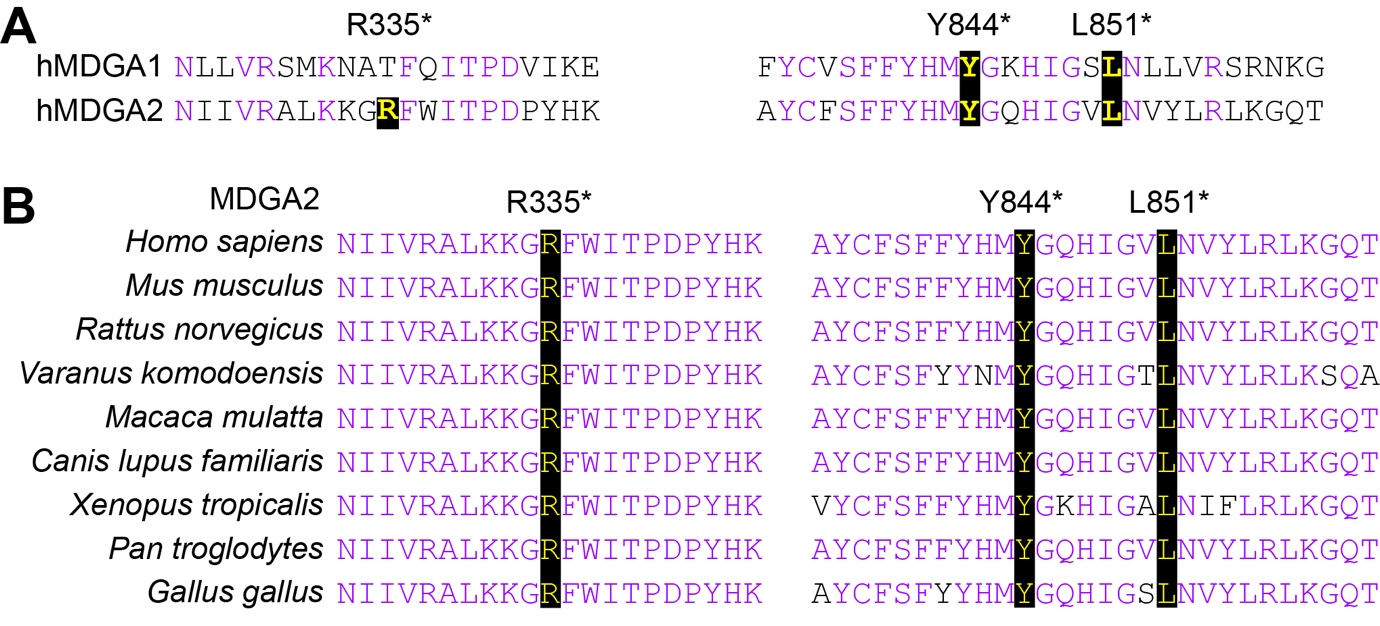

**Figure S8. Alignment and conservation across different species of MDGA2 residues** used in the molecular replacement experiment

(**A**) Alignment of human MDGA1 and MDGA2 amino acids surrounding the mutated residues used in the molecular replacement experiment. The target mutated residues are indicated in bold.

(**B**) Similarity or identity of mutated residues investigated in the current study across different species. The following GenBank accession numbers were utilized for sequence alignment: human MDGA1, NP_705691.1; human MDGA2, NP_001106970.4; mouse MDGA2, NP_001180195.2; rat MDGA2, NP_954890.1; komodo dragon MDGA2, XP_044292213.1; Rhesus monkey MDGA2, XP_001097544.1; dog MDGA2, XP_038529478.1; tropical clawed frog MDGA2, XP_031747317.1; chimpanzee MDGA2, XP_016781528.1; and chicken MDGA2, XP_025006731.1.

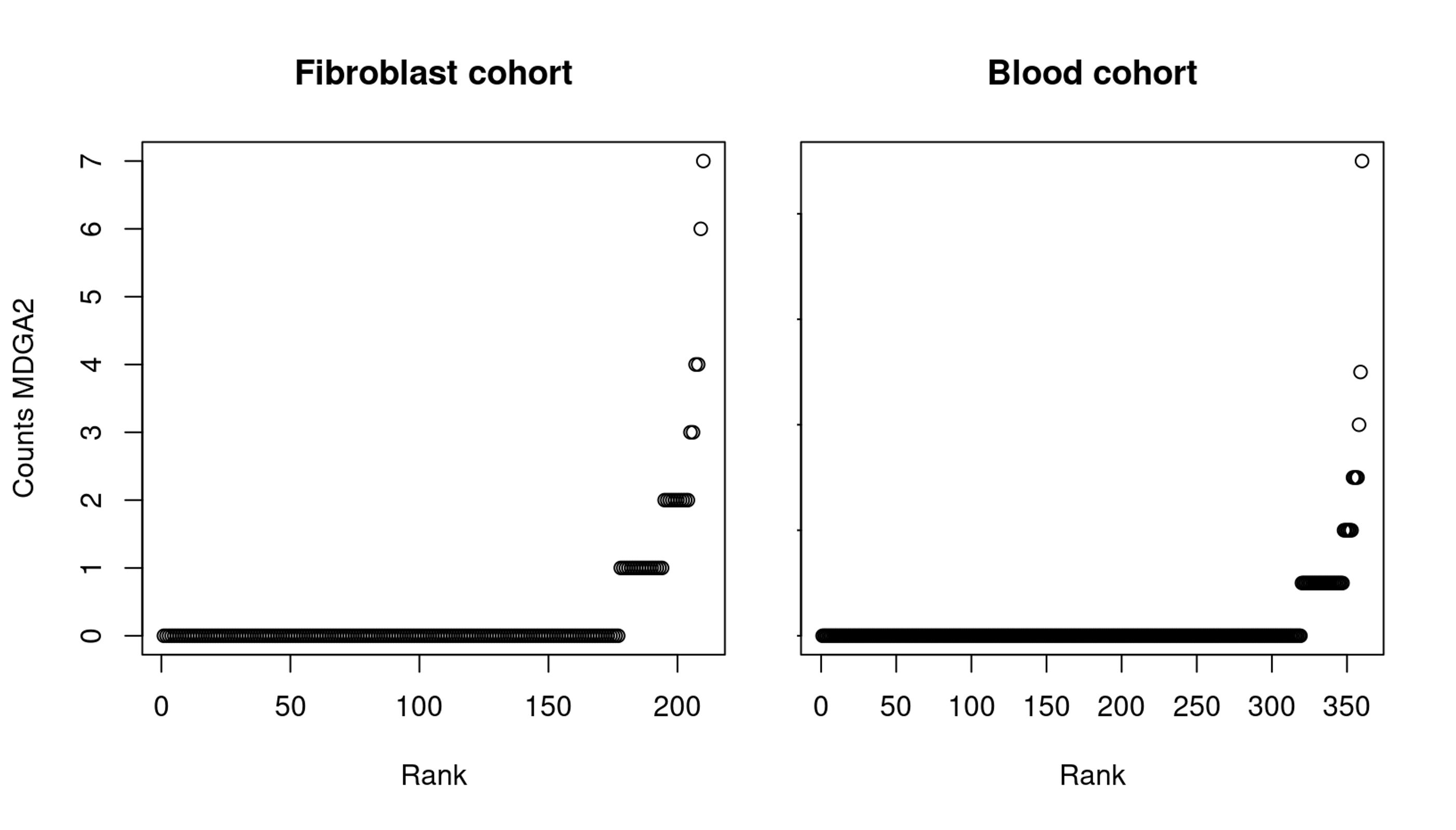

**Figure S9: Sorted raw read counts aligned to *MDGA2* for a cohort composed of blood and fibroblast samples.** To increase sample size, the Solve-RD cohort was used. Most samples have zero or very low counts for both tissues deeming the gene with a too low expression to be properly modelled for expression and splicing analyses.

**Supplemental tables**

**Legends for Excel Tables**

**Table S1: Detailed phenotype of affected individuals with homozygous *MDGA2- related DEE*.**  Clinical description includes parental consanguinity, family history, ethnicity, growth parameters at birth and at last follow up,

developmental milestones, behaviour, sleep, seizures, neurological assessment, dysmorphic facial features and neuroimaging findings.

**Table S2:** **Genome-wide runs of homozygosity detected in P1.** The row

corresponding to the 65.86 Mb ROH overlapping *MDGA2* is bolded. Sequencing data were aligned to Human Genome Reference Build 38 (GRCh38)

| **#Chr** | **Begin** | **End** | **Size (Mb)** | **Nb_variants** | **Percentage_homozygosity** |
| --- | --- | --- | --- | --- | --- |
| **chr1** | 22598043 | 30325556 | 7.73 | 218 | 98.17 |
| **chr1** | 55039774 | 56178164 | 1.14 | 27 | 88.89 |
| **chr1** | 245927813 | 247921496 | 1.99 | 64 | 96.87 |
| **chr2** | 224970 | 8731212 | 8.51 | 74 | 95.95 |
| **chr2** | 181062633 | 214809366 | 33.75 | 432 | 97.45 |
| **chr3** | 38011715 | 56173420 | 18.16 | 537 | 99.63 |
| **chr3** | 75738577 | 140688105 | 64.95 | 762 | 98.56 |
| **chr4** | 6954332 | 8414255 | 1.46 | 51 | 90.20 |
| **chr4** | 41661604 | 45718231 | 4.06 | 36 | 91.67 |
| **chr4** | 78058103 | 79907514 | 1.85 | 37 | 89.19 |
| **chr5** | 67090353 | 71522502 | 4.43 | 26 | 88.46 |
| **chr6** | 5003747 | 24353379 | 19.35 | 148 | 96.62 |
| **chr6** | 97231660 | 99955936 | 2.72 | 26 | 88.46 |
| **chr6** | 105451851 | 116118057 | 10.67 | 113 | 96.46 |
| **chr6** | 148532731 | 165482336 | 16.95 | 269 | 97.77 |
| **chr7** | 31809121 | 40180813 | 8.37 | 128 | 97.66 |
| **chr7** | 48983526 | 76995642 | 28.01 | 311 | 95.18 |
| **chr7** | 130097040 | 131509104 | 1.41 | 30 | 90.00 |
| **chr8** | 57434625 | 119783402 | 62.35 | 409 | 97.07 |
| **chr8** | 141479310 | 144398223 | 2.92 | 248 | 99.19 |
| **chr9** | 7798518 | 18574408 | 10.78 | 59 | 96.61 |
| **chr10** | 102425224 | 103602347 | 1.18 | 45 | 91.11 |
| **chr11** | 47339460 | 49576626 | 2.24 | 72 | 90.28 |
| **chr11** | 64276812 | 65618735 | 1.34 | 118 | 91.53 |
| **chr12** | 28450179 | 40510810 | 12.06 | 179 | 99.44 |
| **chr12** | 40513782 | 52646627 | 12.13 | 328 | 96.65 |
| **chr12** | 79621127 | 80669485 | 1.05 | 42 | 92.86 |
| **chr12** | 104315781 | 113380923 | 9.07 | 137 | 95.62 |
| **chr13** | 101583169 | 103046280 | 1.46 | 48 | 97.92 |
| **chr13** | 111320751 | 114244779 | 2.92 | 75 | 100.00 |
| **chr14** | **29577305** | **95436970** | **65.86** | **942** | **98.51** |
| **chr18** | 64822142 | 80136961 | 15.31 | 99 | 98.99 |
| **chr19** | 15087953 | 16325451 | 1.24 | 52 | 100.00 |
| **chr20** | 4796013 | 6770387 | 1.97 | 36 | 94.44 |
| **chr20** | 34072384 | 35554486 | 1.48 | 75 | 96.00 |
| **chr20** | 45579372 | 46613630 | 1.03 | 53 | 90.57 |
| **chr22** | 31133786 | 32190645 | 1.06 | 37 | 94.59 |
| **chr22** | 43927194 | 45170821 | 1.24 | 35 | 97.14 |

**Table S3:** **Genome-wide runs of homozygosity detected in P2.** The row

corresponding to the 33.16 Mb ROH overlapping *MDGA2* is bolded. Sequencing data were aligned to Human Genome Reference Build 37 (GRCh37)

| **#Chr** | **Begin** | **End** | **Size (Mb)** | **Nb_variants** | **Percentage_homozygosity** |
| --- | --- | --- | --- | --- | --- |
| **chr1** | 866319 | 1986736 | 1.12 | 216 | 99.54 |
| **chr1** | 244624887 | 249110906 | 4.49 | 200 | 96.5 |
| **chr2** | 14372731 | 15731998 | 1.36 | 33 | 90.91 |
| **chr2** | 120221585 | 125405302 | 5.18 | 53 | 90.57 |
| **chr2** | 209051586 | 210809729 | 1.76 | 39 | 100 |
| **chr3** | 55041696 | 56763525 | 1.72 | 29 | 89.66 |
| **chr4** | 100356584 | 103822179 | 3.47 | 32 | 96.87 |
| **chr4** | 103822492 | 141543997 | 37.72 | 311 | 96.14 |
| **chr4** | 163058512 | 183594117 | 20.54 | 130 | 97.69 |
| **chr4** | 187175722 | 188629770 | 1.45 | 27 | 96.3 |
| **chr5** | 64273018 | 67576330 | 3.3 | 42 | 97.62 |
| **chr5** | 70308251 | 71492734 | 1.18 | 29 | 93.1 |
| **chr5** | 96518792 | 102249546 | 5.73 | 26 | 88.46 |
| **chr6** | 34389796 | 91228933 | 56.84 | 907 | 97.91 |
| **chr7** | 2598908 | 5983070 | 3.38 | 85 | 96.47 |
| **chr7** | 5983070 | 45957172 | 39.97 | 627 | 97.61 |
| **chr7** | 50070921 | 51094216 | 1.02 | 33 | 90.91 |
| **chr8** | 131964032 | 139674213 | 7.71 | 131 | 94.66 |
| **chr10** | 3201039 | 5032284 | 1.83 | 28 | 92.86 |
| **chr10** | 114224502 | 116091555 | 1.87 | 64 | 98.44 |
| **chr10** | 129880911 | 135439925 | 5.56 | 173 | 97.11 |
| **chr11** | 130332338 | 132307422 | 1.98 | 36 | 88.89 |
| **chr13** | 48563008 | 68802339 | 20.24 | 115 | 92.17 |
| **chr13** | 85369120 | 103289409 | 17.92 | 121 | 95.87 |
| **chr14** | **39818145** | **72977054** | **33.16** | **460** | **96.74** |
| **chr15** | 83781599 | 85411285 | 1.63 | 41 | 92.68 |
| **chr15** | 85476441 | 86697596 | 1.22 | 45 | 93.33 |
| **chr16** | 81639 | 8829715 | 8.75 | 513 | 98.05 |
| **chr18** | 117236 | 2552885 | 2.44 | 52 | 96.15 |
| **chr18** | 4196981 | 8705583 | 4.51 | 65 | 98.46 |
| **chr22** | 48042316 | 51208255 | 3.17 | 180 | 98.33 |

**Table S4:** **Genome-wide runs of homozygosity detected in P3A.** The row

corresponding to the 24.92 Mb ROH overlapping *MDGA2* is bolded. Sequencing data were aligned to Human GRCh38

| **#Chr** | **Begin** | **End** | **Size(Mb)** | **Nb_variants** | **Percentage_homozygosity** |
| --- | --- | --- | --- | --- | --- |
| **chr1** | 3866195 | 8949347 | 5.08 | 104 | 99.04 |
| **chr2** | 224886539 | 227254745 | 2.37 | 37 | 89.19 |
| **chr3** | 121324510 | 122337585 | 1.01 | 41 | 90.24 |
| **chr3** | 129315915 | 131549980 | 2.23 | 105 | 91.43 |
| **chr6** | 1960101 | 5001040 | 3.04 | 53 | 90.57 |
| **chr6** | 27472225 | 29044183 | 1.57 | 29 | 89.66 |
| **chr7** | 82170712 | 93104453 | 10.93 | 115 | 100.00 |
| **chr7** | 102572565 | 125846926 | 23.27 | 159 | 96.23 |
| **chr9** | 131260861 | 132401397 | 1.14 | 69 | 91.30 |
| **chr10** | 98433922 | 100042505 | 1.61 | 28 | 92.86 |
| **chr10** | 118707701 | 119805347 | 1.10 | 30 | 93.33 |
| **chr11** | 78038571 | 82733048 | 4.69 | 34 | 88.24 |
| **chr13** | 49291591 | 50713272 | 1.42 | 32 | 96.87 |
| **chr13** | 101714392 | 103049340 | 1.33 | 36 | 91.67 |
| **chr14** | **39175294** | **64098864** | **24.92** | **275** | **97.45** |
| **chr14** | 93227076 | 94304653 | 1.08 | 28 | 92.86 |
| **chr16** | 85710126 | 87315927 | 1.61 | 26 | 88.46 |

**Table S5:** **Genome-wide runs of homozygosity detected in P3B.** The row

corresponding to the 18.04 Mb ROH overlapping *MDGA2* is bolded. Sequencing data were aligned to Human GRCh38

| **#Chr** | **Begin** | **End** | **Size(Mb)** | **Nb_variants** | **Percentage_homozygosity** |
| --- | --- | --- | --- | --- | --- |
| **chr2** | 38582020 | 40164829 | 1.58 | 35 | 88.57 |
| **chr2** | 187584169 | 190436642 | 2.85 | 40 | 95.00 |
| **chr3** | 175233966 | 179874452 | 4.64 | 41 | 90.24 |
| **chr4** | 172882763 | 182448903 | 9.57 | 42 | 95.24 |
| **chr5** | 79762994 | 81123530 | 1.36 | 35 | 88.57 |
| **chr5** | 90473830 | 94524094 | 4.05 | 30 | 93.33 |
| **chr6** | 35477434 | 37381257 | 1.90 | 66 | 92.42 |
| **chr7** | 90604304 | 92397873 | 1.79 | 26 | 92.31 |
| **chr10** | 78289930 | 87234695 | 8.94 | 108 | 94.44 |
| **chr10** | 87367434 | 99811771 | 12.44 | 234 | 98.29 |
| **chr10** | 125027777 | 131146695 | 6.12 | 82 | 96.34 |
| **chr14** | **46059348** | **64098864** | **18.04** | **270** | **97.41** |
| **chr14** | 93227076 | 94304653 | 1.08 | 27 | 96.30 |
| **chr16** | 7653783 | 8746097 | 1.09 | 28 | 89.29 |
| **chr16** | 49822780 | 51138766 | 1.32 | 34 | 97.06 |
| **chr16** | 71066935 | 72793859 | 1.73 | 34 | 88.24 |
| **chr18** | 35242388 | 36625305 | 1.38 | 32 | 93.75 |
| **chr20** | 37401743 | 47663405 | 10.26 | 141 | 98.58 |

**Table S6:** **Genome-wide runs of homozygosity detected in P4.** The row

corresponding to the 81.21 Mb ROH overlapping *MDGA2* is bolded. Sequencing data were aligned to Human GRCh38

| **#Chr** | **Begin** | **End** | **Size (Mb)** | **Nb_variants** | **Percentage_homozygosity** |
| --- | --- | --- | --- | --- | --- |
| **chr1** | 28048304 | 34718483 | 6.67 | 122 | 95.08 |
| **chr1** | 86482486 | 91515092 | 5.03 | 63 | 88.89 |
| **chr1** | 91730044 | 94014481 | 2.28 | 37 | 94.59 |
| **chr1** | 210404557 | 248474242 | 38.07 | 588 | 98.47 |
| **chr2** | 16562185 | 34559094 | 18.00 | 262 | 98.09 |
| **chr2** | 65313804 | 79874195 | 14.56 | 134 | 94.78 |
| **chr2** | 111092850 | 113563106 | 2.47 | 77 | 96.10 |
| **chr2** | 208171988 | 209982143 | 1.81 | 32 | 93.75 |
| **chr3** | 3034759 | 4661973 | 1.63 | 28 | 92.86 |
| **chr3** | 34683758 | 56662606 | 21.98 | 567 | 97.71 |
| **chr3** | 57301848 | 58640086 | 1.34 | 72 | 88.89 |
| **chr3** | 68987373 | 155564869 | 86.58 | 977 | 98.87 |
| **chr4** | 91590330 | 99207596 | 7.62 | 56 | 92.86 |
| **chr5** | 110098609 | 115136913 | 5.04 | 58 | 100.00 |
| **chr6** | 350940 | 3723530 | 3.37 | 42 | 100.00 |
| **chr7** | 7531328 | 13688201 | 6.16 | 37 | 94.59 |
| **chr7** | 49775897 | 51026519 | 1.25 | 27 | 92.59 |
| **chr8** | 3493823 | 7414857 | 3.92 | 39 | 94.87 |
| **chr8** | 7841005 | 12174539 | 4.33 | 71 | 94.37 |
| **chr10** | 27037092 | 28534197 | 1.50 | 44 | 90.91 |
| **chr10** | 71439744 | 79304998 | 7.87 | 96 | 97.92 |
| **chr11** | 126446715 | 134374229 | 7.93 | 68 | 98.53 |
| **chr12** | 119857421 | 129081739 | 9.22 | 210 | 98.57 |
| **chr13** | 33126237 | 90362688 | 57.24 | 265 | 97.36 |
| **chr14** | **24413106** | **105624818** | **81.21** | **1177** | **98.56** |
| **chr14** | 105643720 | 106802973 | 1.16 | 92 | 91.30 |
| **chr15** | 96960870 | 101966322 | 5.01 | 85 | 98.82 |
| **chr16** | 24968868 | 27461451 | 2.49 | 30 | 90.00 |
| **chr17** | 137637 | 1477711 | 1.34 | 28 | 96.43 |
| **chr17** | 49312652 | 52636067 | 3.32 | 81 | 96.30 |
| **chr17** | 66732589 | 68269226 | 1.54 | 40 | 90.00 |
| **chr18** | 10789016 | 23714153 | 12.93 | 109 | 94.50 |
| **chr19** | 19934123 | 21505693 | 1.57 | 38 | 92.11 |
| **chr20** | 37168172 | 38350730 | 1.18 | 38 | 89.47 |
| **chr22** | 25874223 | 35720488 | 9.85 | 180 | 98.33 |

**Table S7:** **Genome-wide runs of homozygosity detected in P5A.** The row

corresponding to the 97.14 Mb ROH overlapping *MDGA2* is bolded. Sequencing data were aligned to Human Genome Reference Build 37 (GRCh37)

| **#Chr** | **Begin** | **End** | **Size (Mb)** | **Nb_variants** | **Percentage_homozygosity** |
| --- | --- | --- | --- | --- | --- |
| **chr1** | 17597721 | 23236826 | 5.64 | 121 | 95.87 |
| **chr2** | 85554080 | 87100519 | 1.55 | 34 | 91.18 |
| **chr2** | 208693261 | 210654225 | 1.96 | 34 | 88.24 |
| **chr2** | 215869066 | 217123958 | 1.25 | 29 | 93.1 |
| **chr3** | 105260524 | 108635099 | 3.37 | 27 | 100 |
| **chr4** | 74285239 | 95170839 | 20.89 | 177 | 96.61 |
| **chr6** | 32975381 | 34208614 | 1.23 | 44 | 93.18 |
| **chr8** | 84762193 | 101253092 | 16.49 | 77 | 94.81 |
| **chr11** | 31454975 | 34668175 | 3.21 | 54 | 88.89 |
| **chr11** | 44286566 | 47431966 | 3.15 | 45 | 91.11 |
| **chr11** | 58978684 | 61161357 | 2.18 | 60 | 93.33 |
| **chr13** | 51915371 | 53602970 | 1.69 | 32 | 100 |
| **chr13** | 113720476 | 114792842 | 1.07 | 39 | 92.31 |
| **chr14** | **39818145** | **78285461** | **38.47** | **350** | **97.14** |
| **chr15** | 64017685 | 65756196 | 1.74 | 26 | 96.15 |
| **chr16** | 15116625 | 19548152 | 4.43 | 50 | 92 |
| **chr16** | 65646591 | 68713730 | 3.07 | 59 | 96.61 |
| **chr18** | 29797958 | 46385948 | 16.59 | 55 | 92.73 |
| **chr19** | 11516007 | 13868669 | 2.35 | 38 | 94.74 |
| **chr20** | 11990044 | 25754225 | 13.76 | 113 | 96.46 |
| **chr20** | 29634049 | 39797720 | 10.16 | 101 | 98.02 |
| **chr22** | 26231312 | 35478486 | 9.25 | 66 | 98.48 |

**Table S8:** **Genome-wide runs of homozygosity detected in P5B.** The row

corresponding to the 95.4 Mb ROH overlapping *MDGA2* is bolded. Sequencing data were aligned to Human Genome Reference Build 37 (GRCh37)

| **#Chr** | **Begin** | **End** | **Size (Mb)** | **Nb_variants** | **Percentage_homozygosity** |
| --- | --- | --- | --- | --- | --- |
| **chr1** | 22141030 | 23751140 | 1.61 | 59 | 88.14 |
| **chr1** | 24978892 | 27679797 | 2.7 | 45 | 93.33 |
| **chr1** | 234593403 | 243736210 | 9.14 | 117 | 95.73 |
| **chr2** | 101541798 | 105859249 | 4.32 | 33 | 100 |
| **chr2** | 135102453 | 140655337 | 5.55 | 27 | 88.89 |
| **chr2** | 209030578 | 211302627 | 2.27 | 27 | 92.59 |
| **chr2** | 220356520 | 222321340 | 1.96 | 35 | 91.43 |
| **chr3** | 121228400 | 122354052 | 1.13 | 50 | 92 |
| **chr6** | 46135984 | 48036327 | 1.9 | 53 | 92.45 |
| **chr7** | 540741 | 4965195 | 4.42 | 77 | 96.1 |
| **chr7** | 79833026 | 89847453 | 10.01 | 55 | 92.73 |
| **chr9** | 132597062 | 133914570 | 1.32 | 28 | 92.86 |
| **chr11** | 31454975 | 34668175 | 3.21 | 46 | 91.3 |
| **chr11** | 44639931 | 47431966 | 2.79 | 46 | 95.65 |
| **chr11** | 58978684 | 61161357 | 2.18 | 72 | 95.83 |
| **chr13** | 51915371 | 68802339 | 16.89 | 43 | 88.37 |
| **chr13** | 113742595 | 114792842 | 1.05 | 43 | 88.37 |
| **chr14** | **24976574** | **78285461** | **53.31** | **434** | **95.39** |
| **chr14** | 100759046 | 103167569 | 2.41 | 47 | 91.49 |
| **chr15** | 59515434 | 67782440 | 8.27 | 96 | 95.83 |
| **chr19** | 11516007 | 13868669 | 2.35 | 40 | 95 |
| **chr20** | 30452782 | 35240740 | 4.79 | 71 | 95.77 |
| **chr22** | 22989622 | 28021457 | 5.03 | 69 | 94.2 |

**Table S9:** **Genome-wide runs of homozygosity detected in P6.** The row

corresponding to the 97.45Mb ROH overlapping *MDGA2* is bolded, which is part of a larger slightly interrupted ROH of around282 Mb. Sequencing data were aligned to Human Genome Reference Build 37 (GRCh37)

| **#Chr** | **Begin** | **End** | **Size (Mb)** | **Nb_variants** | **Percentage_homozygosity** |
| --- | --- | --- | --- | --- | --- |
| chr1 | 17707739 | 19181015 | 1.47 | 38 | 89.47 |
| chr1 | 108313258 | 109394593 | 1.08 | 35 | 88.57 |
| chr1 | 112018657 | 114133022 | 2.11 | 70 | 91.43 |
| chr2 | 26152485 | 27324340 | 1.17 | 81 | 88.89 |
| chr2 | 31565316 | 33585993 | 2.02 | 84 | 90.48 |
| chr2 | 38976957 | 39982698 | 1.01 | 50 | 92 |
| chr2 | 140004513 | 141457985 | 1.45 | 27 | 92.59 |
| chr3 | 3136804 | 4716214 | 1.58 | 56 | 89.29 |
| chr3 | 55108358 | 56771523 | 1.66 | 29 | 100 |
| chr3 | 189348968 | 195506542 | 6.16 | 210 | 93.33 |
| chr3 | 195510683 | 197762623 | 2.25 | 124 | 95.97 |
| chr4 | 38698924 | 62694717 | 24 | 406 | 93.6 |
| chr4 | 79462311 | 101953322 | 22.49 | 387 | 92.76 |
| chr4 | 119772220 | 121550059 | 1.78 | 28 | 89.29 |
| chr5 | 95124328 | 96430832 | 1.31 | 57 | 89.47 |
| chr5 | 118183609 | 121488506 | 3.3 | 34 | 91.18 |
| chr5 | 170640827 | 175394181 | 4.75 | 53 | 88.68 |
| chr5 | 176069864 | 180687428 | 4.62 | 330 | 96.06 |
| chr6 | 34069822 | 35203203 | 1.13 | 28 | 89.29 |
| chr6 | 44221145 | 45922849 | 1.7 | 60 | 96.67 |
| chr6 | 86259340 | 88757663 | 2.5 | 42 | 90.48 |
| chr6 | 146480740 | 152456475 | 5.98 | 131 | 94.66 |
| chr6 | 152884188 | 154478440 | 1.59 | 29 | 89.66 |
| chr7 | 40356554 | 44043357 | 3.69 | 30 | 93.33 |
| chr7 | 44102777 | 54103946 | 10 | 203 | 98.03 |
| chr8 | 126445544 | 143352528 | 16.91 | 320 | 96.56 |
| chr9 | 127781259 | 129143435 | 1.36 | 27 | 88.89 |
| chr10 | 3155698 | 4879568 | 1.72 | 117 | 94.87 |
| chr12 | 29908580 | 32890685 | 2.98 | 125 | 96.8 |
| chr12 | 65114065 | 66725160 | 1.61 | 33 | 96.97 |
| chr12 | 75692607 | 76740149 | 1.05 | 32 | 90.62 |
| chr12 | 97921533 | 104126600 | 6.21 | 105 | 89.52 |
| chr12 | 104131384 | 110924566 | 6.79 | 216 | 94.44 |
| chr14 | 21861133 | 35577597 | 13.72 | 487 | 94.05 |
| chr14 | 35593477 | 44974137 | 9.38 | 90 | 91.11 |
| **chr14** | **46315127** | **51372098** | **5.06** | **157** | **97.45** |
| chr14 | 60032046 | 61113325 | 1.08 | 51 | 90.2 |
| chr15 | 66249763 | 67476952 | 1.23 | 79 | 94.94 |
| chr15 | 81217848 | 86768135 | 5.55 | 157 | 93.63 |
| chr15 | 99548905 | 100881214 | 1.33 | 49 | 95.92 |
| chr16 | 5037498 | 6145753 | 1.11 | 41 | 90.24 |
| chr16 | 7704022 | 8862956 | 1.16 | 42 | 95.24 |
| chr16 | 81254052 | 83940480 | 2.69 | 77 | 94.81 |
| chr17 | 10223697 | 11532835 | 1.31 | 112 | 97.32 |
| chr18 | 3253543 | 14542890 | 11.29 | 298 | 96.64 |
| chr18 | 14780130 | 22008913 | 7.23 | 104 | 98.08 |
| chr18 | 27598866 | 55915969 | 28.32 | 428 | 93.69 |
| chr18 | 56648756 | 70532185 | 13.88 | 235 | 97.02 |
| chr19 | 536107 | 4511283 | 3.98 | 870 | 96.78 |
| chr19 | 4511581 | 5787214 | 1.28 | 95 | 96.84 |
| chr19 | 5787234 | 7970635 | 2.18 | 323 | 95.98 |
| chr20 | 55929881 | 62571417 | 6.64 | 479 | 95.82 |
| chr22 | 17072483 | 21403375 | 4.33 | 308 | 96.1 |
| chr22 | 21742684 | 24579049 | 2.84 | 206 | 97.09 |
| chr22 | 25024503 | 28021457 | 3 | 77 | 97.4 |

**Materials and Methods**

**Identification and recruitment of affected individuals**

Participants were recruited through SYNAPS project, a global collaborative project to unravel genetic causes of rare neurological or neurodevelopmental disorders. Two additional families were then identified through Gene Matcher^1^ (F2, F5). Informed consent for genetic analyses was obtained from all subjects. Parents and legal guardians of all affected individuals gave their consent for the publication of clinical and genetic information according to the Declaration of Helsinki, and the study was approved by the Research Ethics Committee, Institute of Neurology, University College London (ION UCL) (07/Q0512/26) and the local Ethics Committees of each participating centre. All clinical data and family history were collected, and family pedigrees were constructed using Quickped shiny app.^2^

Proband-only whole exome sequencing (WES) was performed for all families. WES libraries were prepared using a SureSelect All Exon V6 kit (Agilent) and sequenced on Illumina platforms, as described before, in Macrogen or at collaborating centers. Variant annotation was performed with the Ensembl Variant Effect Predictor v.103,^3^ incorporating allele frequencies from multiple publicly available population databases (including gnomAD v.3.1.2 (https://gnomad.broadinstitute.org/) and in-house research data (Queen Square Genomics and UCL SYNaPS Study Group), where only rare variants (Minor allele Frequency less than 0.01) were retained. The bioinformatics filtering strategy included screening for only exonic and donor/acceptor splicing variants. Variant files were analysed for runs of homozygosity (ROH) using Automap v1.0.^4^ All identified MDGA2 variants were described according to the recommendations of the Human Genome Variation Society and are based on reference sequence GenBank: NM_001113498.3.

**Expression vectors.** pDisplay-MDGA2 nonsense variants (R335*, Y844*, and L851*) were generated by polymerase chain reaction (PCR)-based mutagenesis, which was performed using PrimeSTAR HS DNA polymerase (Takara) and a pDisplay MDGA2 wild-type (WT) construct as a template. L-313 MDGA2 variants (R335*, Y844*, and L851*) were constructed by PCR amplifying the indicated full-length MDGA2 variants, digesting them with *Nhe*I and *BsrG*I, and subcloning the resulting products into the L-313 vector ^5^. The following constructs were as described previously: pDisplay-Nlgn1 WT and pCMV-IgC Nlgn1.^6^

**Antibodies.** The following commercially available antibodies were used: mouse monoclonal anti-β-actin (clone AC-74; Sigma-Aldrich; Cat# sc-47778; RRID: AB_476743); mouse monoclonal anti-Nlgn1 (Synaptic Systems; Cat# 129 111; RRID: AB_887747); guinea pig polyclonal anti-VGLUT1 (Millipore; Cat# ab5905; RRID: AB_2301751); mouse monoclonal anti-PSD-95 (clone K28/43, Neuromab; Cat# 75-028; RRID: AB_2877189); goat polyclonal anti-EGFP (Rockland; Cat# 600-101-215; RRID: AB_218182); mouse monoclonal anti-HA (clone 16B12; BioLegend; Cat# 901501; RRID: AB_2565006); Cy3-donkey anti-human IgG antibodies (Jackson ImmunoResearch; Cat# 709-165-149; RRID: AB_2340535); Cy3-AffiniPure donkey Anti-rabbit IgG antibodies (Jackson ImmunoResearch; Cat# 711-165-152; RRID: AB_2307443); Cy3-AffiniPure donkey anti-mouse IgG antibodies (Jackson ImmunoResearch; Cat# 715-165-150; RRID: AB_2340813); Cy3-donkey anti-human IgG antibodies (Jackson ImmunoResearch; Cat# 709-165-149; RRID: AB_2340535); Cy3-donkey anti-guinea pig IgG antibodies (Jackson ImmunoResearch; Cat# 706-035-148; RRID: AB_ 2340447); FITC-AffiniPure donkey anti-mouse IgG antibodies (Jackson ImmunoResearch; Cat# 715-095-150; RRID: AB_2340792) ; FITC-AffiniPure donkey anti-goat IgG antibodies (Jackson ImmunoResearch; Cat# 705-095-147; RRID: AB_2340401); and FITC-AffiniPure Donkey anti-rabbit IgG antibodies (Jackson ImmunoResearch; Cat# 711-095-152; RRID: AB_2315776).

**Cell culture.** HEK293T cells (ATCC; CRL-3216) were cultured in Dulbecco’s modified Eagle’s medium (DMEM; Welgene) supplemented with 10% fetal bovine serum (FBS; Tissue Culture Biologicals) and 1% penicillin-streptomycin (Thermo Fisher) at 37 °C in a humidified 5% CO_2_ atmosphere. All procedures were performed according to the guidelines and protocols for rodent experimentation approved by the Institutional Animal Care and Use Committee of Daegu Gyeongbuk Institute of Science and Technology (DGIST).

**Preparation of lentiviruses.** Recombinant lentiviruses were produced as previously described ^7^. In brief, HEK293T cells were transfected at a 2:2:1 ratio with a lentiviral vector, psPAX2, and pMD2G, using polyethylenimine (PEI) according to the manufacturer’s protocol. At 72 h post-transfection, lentiviruses were harvested by collecting the conditioned medium and subjecting it to a brief centrifugation at 1,000 × g to remove cellular debris. Filtered media containing 5% sucrose were centrifuged at 117,969 × g for 2 h, supernatants were removed, and each viral pellet was washed with ice-cold PBS and resuspended in 80 μl PBS.

**Transfection, immunocytochemistry, confocal microscopy imaging, and analyses.** Hippocampal cultured neurons were prepared from E18 rat brains and cultured on poly-L-lysine-coated coverslips in Neurobasal medium supplemented with B-27 (ThermoFisher Scientific), 0.5% FBS, 0.5 mM GlutaMax (ThermoFisher Scientific), and sodium pyruvate (ThermoFisher Scientific), as previously described. ^8^ Cultured neurons at DIV5 were cotransfected with vectors encoding EGFP and indicated MDGA1 using a CalPhos Transfection Kit (Takara), and immunostained at DIV14. For immunocytochemistry, cultured neurons were fixed with 4% paraformaldehyde/4% sucrose, permeabilized with 0.2% Triton X-100 in phosphate buffered saline (PBS), immunostained with the indicated primary antibodies, and detected with the indicated Cy3-, fluorescein isothiocyanate (FITC)-, or Alexa Fluor 647-conjugated secondary antibodies (Jackson ImmunoResearch). Transfected neurons were chosen randomly, and images were acquired using a confocal microscope (LSM800; Zeiss) with a 63 × objective lens; all image settings were kept constant. Z stack images were converted to maximal intensity projections and analyzed to obtain the size, intensity, and density of puncta immunoreactivities derived from marker proteins. Quantification was performed in a blinded manner using the MetaMorph software (Molecular Devices).

**Electrophysiolical analyses of cultured hippocampal neurons.** Cultured hippocampal neurons were prepared as previously described ^6^. Neurons obtained from WT mice were transfected with vectors encoding the indicated MDGA2 variant and recorded and analyzed at DIV13–16. Using a Model P-97 pipette puller (RWD Life Science), pipettes were pulled from borosilicate glass (i.d., 0.86 mm; o.d., 1.5 mm; Sutter Instruments). The resistances of internal solution-containing patch pipettes ranged from 3 to 6MΩ. The utilized internal solutions for each recording type were as follows: 145 mM CsCl, 0.3 mM Na-GTP, 10 mM EGTA, 10 mM HEPES, 5 mM NaCl, and 4 mM Mg-ATP, with pH adjusted to 7.2 to 7.4 with CsOH and an osmolarity of 290 to 295 mOsmol/L for mEPSCs; and 130 mM Cs-MeSO_4_, 5 mM TEA-Cl, 1 mM QX-314, 0.5 mM EGTA, 8 mM NaCl, 10 mM HEPES, 0.4 mM GTP-Na, 10 mM phosphocreatine-Na2, and 4 mM ATP-Mg, with pH adjusted to 7.2 to 7.4 with CsOH and an osmolarity of 290 to 295 mOsmol/L for recording evoked NMDA and AMPA EPSC-paired pulse ratios (PPRs). The external solution included 130 mM NaCl,1 mM MgCl2, 2 mM CaCl2, 4 mM KCl, 10 mM D-glucose, and 10 mM HEPES, with pH adjusted to 7.2 to 7.4 with NaOH and an osmolarity of 300 to 305 mOsmol/L. The whole-cell arrangement was built at room temperature using M-TSC manipulators (SENSAPEX). Electrophysiological data were obtained using an Axon Multiclamp 700B amplifier and the pCLAMP software and digitized using an Axon DigiData 1550B data capture board (Axon Instruments). mEPSCs and evoked AMPA-EPSCs were measured at a holding potential of −70 mV and evoked NMDA-EPSCs were measured at a holding potential of +40 mV. Synaptic currents were analyzed offline using the Clampfit 10.8 software (Molecular Devices). For recording of mEPSCs, the external solution was supplemented with 1 μM TTX and 50 μM picrotoxin to block Na^+^ currents and GABA_A_ receptors, respectively. For recording of evoked NMDA EPSCs, 10 µM picrotoxin and 10 µM CNQX were added to the extracellular solution. For recording of evoked AMPA EPSCs, 10 µM picrotoxin and 50 µM D-AP5 were added to the extracellular solution. For measurement of PPRs, evoked synaptic responses were stimulated (0.2 ms-current pulses) using a bipolar concentric electrode positioned 200 to 300 mm in front of postsynaptic pyramidal neurons at intensities producing 40 to 50% of the maximal EPSCs. The drugs required for electrophysiological recordings were purchased as follows: 6-cyano-7-nitroquinoxaline-2,3-dione (CNQX) from Sigma-Aldrich (Cat #C127), picrotoxin from Tocris (Cat #1128), tetrodoxin (TTX) from Tocris (Cat #1069), and D-APV from Tocris (Cat #0106).

**Cell-surface binding assays.** Ig-fused Nlgn1 and IgC alone (control) were produced in HEK293T cells. Soluble Ig-fused Nlgn1 was purified using protein A-Sepharose beads (GE Healthcare) as previously described. ^9^ Bound proteins were eluted with 0.1 M glycine (pH 2.5) and immediately neutralized with 1 M Tris-HCl (pH 8.0). Transfected HEK293T cells expressing the indicated plasmids were incubated with 10 μg/ml Ig-fused proteins for 2 h at 37°C with gentle agitation. Images were acquired using a confocal microscope (LSM800; Carl Zeiss).

**Staining for surface/intracellular proteins.** HEK293T cells were transfected with expression vectors encoding HA-MDGA2 WT or the indicated variants. After 48 h, cells were washed twice with PBS, fixed with 3.7% formaldehyde for 10 min at 4°C, and blocked with 3% horse serum/0.1% bovine serum albumin (BSA; crystalline grade) in PBS for 15 min at room temperature. Surface-expressed proteins were stained with mouse anti-HA antibody at room temperature. After 90 min, the cells were washed twice with PBS and incubated with Cy3-conjugated anti-mouse antibodies for 1 h at room temperature. The cells were then permeabilized with PBS containing 0.2% Triton X-100 for 10 min at 4°C, incubated with rabbit anti-HA antibody for 90 min at room temperature to label intracellular proteins, and incubated with FITC-conjugated anti-rabbit secondary antibodies. Images were acquired using a confocal microscope (LSM800; Carl Zeiss).

**Heterologous synapse formation assays.** HEK293T cells were transfected with vectors encoding EGFP (negative control), the indicated MDGA2 variants, or Nlgn1, using polyethylenimine as previously described ^7^. After 48 h, the transfected HEK293T cells were trypsinized, seeded onto hippocampal neurons at DIV12, cocultured for an additional 24 h, and double-immunostained at DIV13 with antibodies against EGFP, HA, and synapsin. All images were acquired using a confocal microscope (LSM800; Zeiss). For quantification, the contours of transfected HEK293T cells were chosen as the region of interest (ROI). Fluorescence intensities of synaptic markers in each ROI were quantified for the red and green channels using the MetaMorph software (Molecular Devices). The normalized synapse density for transfected HEK293T cells was expressed as the ratio of red to green fluorescence.

**Quantification and statistical analysis.** Data analysis and statistical tests were performed using the GraphPad Prism7.0 software (RRID: SCR_002798). Heterologous synapse-formation assays and surface-binding assay results were quantified by identifying randomly selecting transfected HEK293T cells as the ROI. The fluorescence intensities of synaptic marker puncta or Fc-fusion proteins were normalized to transfected protein signal intensities using the MetaMorph software (Molecular Devices). All data are expressed as means ± standard error of the mean (SEM) unless stated otherwise, and significance is indicated with an asterisk. All experiments were performed using at least three cultures, and the normality of data distributions was evaluated using the Shapiro-Wilk test. Data were compared using Student’s t-test or one-way analysis of variance (ANOVA), with a non-parametric Kruskal-Wallis test followed by Dunn’s multiple comparison test for post-hoc group comparisons; Mann Whitney *U* test; or Fisher’s least significance difference. The ‘n’ values used are indicated in the figure legends, and the numbers shown indicate replicates. Tests used to determine statistical significance are stated in the related text and figure legends. A *p*-value < 0.05 was considered statistically significant, and individual *p*-values are indicated in the respective figure legends.

**Graphical abstract**

The graphical abstract was partially created using icons (as is, or modified) from

BioRender. Ramakrishna, N. (2025) https://BioRender.com/h26g880. Other icons

were made on Adobe Illustrator.
